# Diverse Functional Autoantibodies in Patients with COVID-19

**DOI:** 10.1101/2020.12.10.20247205

**Authors:** Eric Y. Wang, Tianyang Mao, Jon Klein, Yile Dai, John D. Huck, Feimei Liu, Neil S. Zheng, Ting Zhou, Benjamin Israelow, Patrick Wong, Carolina Lucas, Julio Silva, Ji Eun Oh, Eric Song, Emily S. Perotti, Suzanne Fischer, Melissa Campbell, John B. Fournier, Anne L. Wyllie, Chantal B. F. Vogels, Isabel M. Ott, Chaney C. Kalinich, Mary E. Petrone, Anne E. Watkins, Yale IMPACT Team, Charles Dela Cruz, Shelli F. Farhadian, Wade L. Schulz, Nathan D. Grubaugh, Albert I. Ko, Akiko Iwasaki, Aaron M. Ring

**Affiliations:** Department of Immunobiology, Yale School of Medicine, New Haven, CT, USA; Department of Pharmacology, Yale School of Medicine, New Haven, CT, USA; Department of Epidemiology of Microbial Diseases, Yale School of Public Health, New Haven, CT, USA; Department of Medicine, Section of Pulmonary and Critical Care Medicine, Yale School of Medicine, New Haven, CT, USA; Department of Internal Medicine (Infectious Diseases), Yale School of Medicine, New Haven, CT, USA; Department of Laboratory Medicine, Yale School of Medicine, New Haven, CT, USA; Center for Outcomes Research and Evaluation, Yale-New Haven Hospital, New Haven, CT, USA; Howard Hughes Medical Institute, Chevy Chase, MD, USA

**Author notes:** These authors contributed equally to this work (Eric Y. Wang, Tianyang Mao, Jon Klein, Yile Dai). A list of authors and their affiliations appears at the end of the paper. Correspondence (A.M.R.); (A.I.).

## Abstract

COVID-19 manifests with a wide spectrum of clinical phenotypes that are characterized by exaggerated and misdirected host immune responses^1–8^. While pathological innate immune activation is well documented in severe disease^1^, the impact of autoantibodies on disease progression is less defined. Here, we used a high-throughput autoantibody discovery technique called Rapid Extracellular Antigen Profiling (REAP) to screen a cohort of 194 SARS-CoV-2 infected COVID-19 patients and healthcare workers for autoantibodies against 2,770 extracellular and secreted proteins (the “exoproteome”). We found that COVID-19 patients exhibit dramatic increases in autoantibody reactivities compared to uninfected controls, with a high prevalence of autoantibodies against immunomodulatory proteins including cytokines, chemokines, complement components, and cell surface proteins. We established that these autoantibodies perturb immune function and impair virological control by inhibiting immunoreceptor signaling and by altering peripheral immune cell composition, and found that murine surrogates of these autoantibodies exacerbate disease severity in a mouse model of SARS-CoV-2 infection. Analysis of autoantibodies against tissue-associated antigens revealed associations with specific clinical characteristics and disease severity. In summary, these findings implicate a pathological role for exoproteome-directed autoantibodies in COVID-19 with diverse impacts on immune functionality and associations with clinical outcomes.

---

Humoral immunity plays dichotomous roles in COVID-19. Although neutralizing antibodies afford protection against SARS-CoV-2 infection^9,10^, growing evidence suggests that dysregulated humoral immunity also contributes to the characteristic immunopathology of COVID-19^11–17^. For example, subsets of COVID-19 patients commonly exhibit an expansion of pathological extrafollicular B cell populations (IgD^-^ /CD27^-^ double negative, DN) that have been associated with autoantibody production in systemic lupus erythematosus (SLE) patients^12,18^. Furthermore, recent reports have identified isolated autoantibody reactivities in COVID-19 patients, including those that are characteristic of systemic autoimmune diseases such as antinuclear antibodies, rheumatoid factor (anti-IgG-Fc antibodies), antiphospholipid antibodies, and antibodies against type 1 interferons (IFN-I)^15–17^. Importantly, some autoantibodies, particularly neutralizing antibodies against IFN-I, appear to directly contribute to COVID-19 pathophysiology by antagonizing innate antiviral responses^11^. While striking examples of disease-modifying autoantibody responses have been described, the full breadth of autoantibody reactivities in COVID-19 and their immunological and clinical impacts remain undetermined at a proteome-scale. We therefore sought to identify functional autoantibody responses in COVID-19 patients by screening for autoantibody reactivities against the human exoproteome (the set of extracellular and secreted proteins in the proteome).

## COVID-19 patients have widespread autoantibody reactivity against extracellular antigens

To discover functional autoantibodies that could influence COVID-19 outcomes, we used a high-throughput autoantibody discovery method called Rapid Extracellular Antigen Profiling (REAP; Wang et al, manuscript in preparation). REAP enables highly multiplexed detection of antibody reactivities against a genetically-barcoded library of 2,770 human extracellular proteins displayed on the surface of yeast. Briefly, the REAP process involves biopanning of serum/plasma-derived patient IgG against this library, magnetic selection of the IgG-bound clones, and sequencing of the barcodes of the isolated yeast **(Fig. 1a)**. REAP thus converts an antibody:antigen binding event into a quantitative sequencing readout (“REAP Score”) based on the enrichment of each protein’s barcodes before and after selection (see methods). To allow for detection of antibodies against coronavirus proteins, we additionally included the receptor binding domain (RBD) of SARS-CoV-2 and other common coronaviruses in the library (full antigen list in **Supplementary Table 1**). We used REAP to screen samples from people with SARS-CoV-2 infection who were prospectively followed as part of the Yale Implementing Medical and Public Health Action Against Coronavirus CT (IMPACT) study. This cohort includes 172 patients seen at Yale-New Haven Hospital with a range of clinical severities and 22 healthcare workers with mild illness or asymptomatic infection. Longitudinal samples were screened for a subset of the cohort. Patients were excluded from subsequent analysis if they were undergoing active chemotherapy for malignancy; possessed any metastatic disease burden; were receiving pharmacological immunosuppression for solid organ transplant; or had received convalescent COVID-19 plasma as part of a clinical trial. IMPACT patients were next stratified according to COVID-19 disease severity as reported previously^1^ and described briefly in Methods. As healthy controls, we screened 30 healthcare workers who tested SARS-CoV-2-negative by RT-qPCR throughout their follow-up period in the IMPACT study. We concomitantly assessed nasopharyngeal viral RNA load, plasma cytokine/chemokine profiles through a Luminex panel, and blood leukocyte composition by flow cytometry as previously reported^1^. Patient demographics can be found in **Extended Data Table 1**.

**Figure 1:**
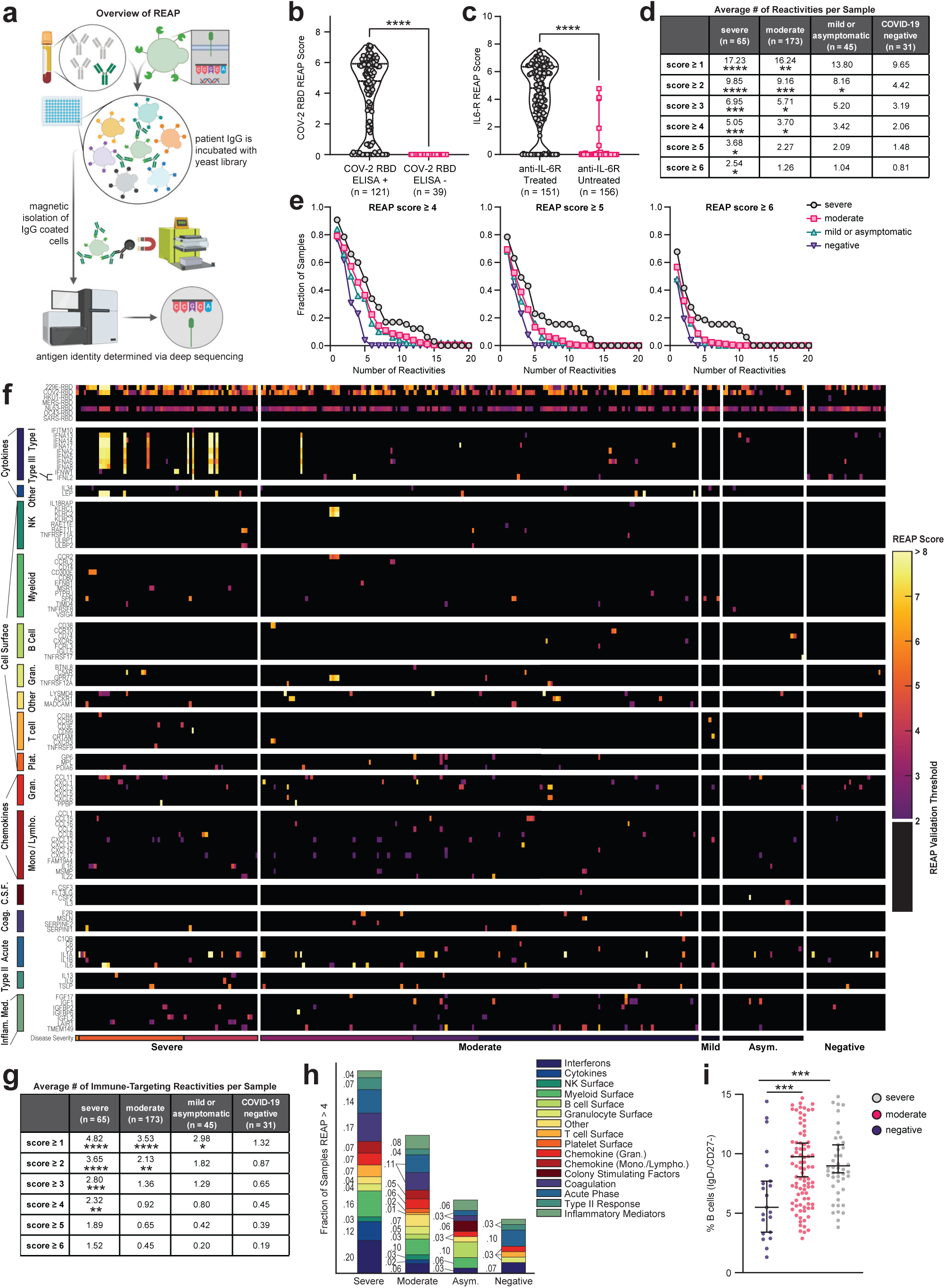
COVID-19 patients have widespread autoantibody reactivity against extracellular antigens. **a**, Simplified schematic of REAP. Antibodies are incubated with a barcoded yeast library displaying members of the exoproteome. Antibody bound yeast are enriched by magnetic column-based sorting and enrichment is quantified by next-generation sequencing. **b**, COV-2 RBD REAP scores for COVID-19 patient samples stratified by positive (n = 121) or negative (n = 39) ELISA RBD reactivity. Significance was determined using a two-sided Mann-Whitney U test. **c**, IL6-R REAP scores for COVID-19 patient samples stratified by treatment with an anti-IL-6R biologic therapy (tocilizumab or sarilumab). Samples collected at least one day after infusion were considered treated. Samples collected on the day of infusion were excluded from analysis due to uncertainty in the timing of sample collection. Significance was determined using a two-sided Mann-Whitney U test. **d**, Average number of positive reactivities per sample at different score cutoffs, stratified by disease severity. A positive reactivity was defined as one with a REAP score greater than or equal to the corresponding score cutoff. Comparisons were made between each disease severity group and the COVID-19 negative group. Significance was determined using a Kruskal-Wallis test followed by a Dunnet’s test. **e**, Distributions of hits between samples of different disease severities at different score cutoffs. Each point on the graph represents the fraction of samples in a given severity group that had at least the indicated number of reactivities at the given score cutoff. **f**, Heatmap of immune-related protein REAP scores stratified by disease severity. Scores below the REAP Validation Threshold of 2.0 were set to 0 to aid interpretation of significant hits. **g**, Average number of positive immune-targeting reactivities per sample at different score cutoffs, stratified by disease severity. Analysis was performed as in **d. h**, Fraction of samples, stratified by disease severity, with a REAP score greater than 4 for at least one antigen in each given antigen group. **i**, Percentages of IgD-/CD27-B cells among peripheral leukocytes in patient samples, stratified by disease severity. Significance was determined using a Kruskal-Wallis test followed by a Dunn’s test. Longitudinal samples from the same patient were included in all analyses in this figure. *P ≤; 0.05, **P ≤; 0.01, ***P ≤; 0.001, ****P ≤; 0.0001.

To validate the performance of the REAP platform in this patient cohort, we assessed the concordance of REAP data with known antibody reactivities. We assessed levels of antibody reactivity against SARS-CoV-2 RBD by ELISA for 160 subject samples and compared the results against RBD reactivity as detected by REAP. As seen in **Fig. 1b**, all samples that were ELISA negative for SARS-CoV-2 RBD reactivity were also REAP negative (score = 0), whereas 84% of samples that were ELISA positive were REAP positive (score > 0). Additionally, 151 samples were from patients who received tocilizumab or sarilumab (anti-IL-6R antibodies), which provided an inherent “spike-in” experimental control to further validate REAP. Analysis of IL-6R reactivities by REAP showed strong IL-6R reactivity in these patients compared to those that did not receive anti-IL-6R therapy **(Fig. 1c)**.

Next, we examined the total degree of autoreactivity stratified by COVID-19 disease severity by quantifying the number of autoantibody reactivities at different REAP score thresholds. Irrespective of the REAP score cutoff used, COVID-19 samples had a greater number of reactivities compared to control samples, with the number of reactivities positively correlating with disease severity **(Fig. 1d)**. At score cutoffs 4, 5, and 6, there was a clear difference between severe and moderate/mild COVID-19 samples; the highest scoring reactivities were preferentially enriched in the severe patients **(Fig. 1d,e)**. Of note, there was not a statistically significant difference in days from symptom onset (DFSO) between severe and moderate COVID-19 samples (**Supplementary Fig. 1a**), suggesting that differences in autoantibody reactivities between these two groups were not due to temporal confounding. DFSO data was not available for mild and asymptomatic COVID-19 samples. Compared to REAP profiles of SLE and autoimmune polyendocrinopathy-candidiasis-ectodermal dystrophy (APECED) patients, COVID-19 samples had greater numbers of reactivities compared to SLE, but fewer numbers of reactivities compared to APECED **(Supplementary Fig. 1b)**. Altogether, these results suggest that broad autoreactivity toward the exoproteome is a highly prevalent feature of COVID-19 patients.

To investigate the temporal nature of these reactivities relative to COVID-19, we assessed REAP scores longitudinally for a subset of infected patients. Additionally, given that these samples were collected during the primary wave of infections at our study site from March through May, we considered the probability of re-infection in any patient an unlikely confounder for our temporal analysis. Although definitive assignment was not possible due to a lack of recent pre-infection baseline samples for most patients, we inferred reactivities as likely pre-existing, newly acquired, or waning based on their REAP score trajectories plotted against reported days from symptom onset **(Supplementary Fig. 2)**. For example, approximately 50% of REAP reactivities with a score of 3 or above were present within 10 days from symptom onset, suggesting that they were likely pre-existing **(Supplementary Fig. 2a)**. Around 10% of longitudinal REAP reactivities started with a score of 0 and had an increase in score of at least 1 at a later time point, indicating they were newly acquired post-infection **(Supplementary Fig. 2b)**. Finally, approximately 15% of longitudinal REAP reactivities started with a score of 3 and had a decrease in score of at least 1 at a later time point, which suggests waning antibody titers **(Supplementary Fig. 2c)**. Representative plots of these reactivities are depicted in **Supplementary Fig. 2d-l**.

In order to further explore potential cellular sources of the elevated autoantibody reactivities in COVID-19 patients, we examined B cell phenotypes in peripheral blood mononuclear cells (PBMC) matching the REAP plasma samples. Similar to previous reports, we find that extrafollicular DN B cells are expanded in moderate and severe COVID-19 patients compared to uninfected controls **(Fig. 1i)**.

## Autoantibodies in COVID-19 patients target a wide range of immune-related proteins

Having established that COVID-19 patients have increased autoantibody reactivities against extracellular antigens, we sought to specifically investigate autoantibodies that could impact immune responses (immune-targeting autoantibodies). To this end, we evaluated a group of immunomodulatory antigens with significant differences in REAP scores between symptomatic and negative cohorts and grouped these antigens by their known immunological function and/or association with specific cell types **(Fig. 1f)**. We found that autoantibodies in COVID-19 patients targeted proteins involved in diverse immunological functions such as acute phase response, type II immunity, leukocyte trafficking, interferon responses, and lymphocyte function/activation. Cytokine autoantibody targets included type 1 interferons, IL-1α/β, IL-6, GM-CSF (*CSF2*), IL-18Rβ (*IL18RAP*), and Leptin (*LEP*). Chemokine autoantibody targets included CXCL1, CXCL7 (*PPBP*), CCL2, CCL15, CCL16, and the chemokine decoy receptor ACKR1 (Duffy blood group antigen). Immunomodulatory cell surface autoantibody targets included NKG2D ligands (*RAET1E/L, ULBP1/2*), NK cell receptors NKG2A/C/E (*KLRC1/2/3*), B cell expressed proteins (*CD38, FCMR, FCRL3, CXCR5*), T cell expressed proteins (*CD3E, CXCR3, CCR4*), and myeloid expressed proteins (*CCR2, CD300E*). Stratifying samples by disease severity, we found that immune-targeting autoantibodies were elevated in COVID-19 patients compared to controls and that the fraction of samples with these reactivities increased with worsening disease severity **(Fig. 1g,h)**. Using ELISA, we orthogonally validated a subset of 16 autoantibodies that target cytokines, chemokines, growth factors, complement factors, and cell surface proteins **(Supplementary Fig. 3)**. These results demonstrate that COVID-19 patients possess autoantibodies that may impact a wide range of immunological functions.

## Autoantibodies targeting cytokines/chemokines are associated with distinct virological and immunological characteristics in COVID-19

We hypothesized that the presence of autoantibodies in patients influences the circulating concentrations of their autoantigen targets. We therefore compared the plasma concentrations of cytokines and chemokines measured by Luminex in patients who possessed or lacked autoantibodies against these targets. In some cases, autoantibodies were associated with apparent increases in their autoantigen targets (*e*.*g*., CCL15, CXCL1, IFN-α2, IL-13; **Supplementary Fig. 4b,f,j,m**), whereas in other cases they correlated with apparent decreases (*e*.*g*., IL-1A, IL-1B; **Supplementary Fig. 4k,l**). This is consistent with the ability of antibodies to both neutralize, but also paradoxically to pharmacokinetically stabilize their targets by preventing renal clearance and/or receptor-mediated endocytosis^19^. A caveat to these findings is that autoantibodies can variably interfere with antibody-based detection methods such as the Luminex assay or ELISA, for instance, by blocking antigen capture or detection with secondary reagents. This may explain the apparent discrepancy between our results showing increased circulating IFN-α2 concentrations (though not IFN activity) in antibody-positive patients compared to the findings of Bastard et al.^11^ that used a different assay and found decreased apparent concentrations. Nevertheless, these results provide orthogonal clinical validation that the presence of autoantibodies affected circulating levels of their targets in patients.

To more directly assess potential immunomodulatory effects of cytokine/chemokine targeting autoantibodies in COVID-19 patients, we assessed the *in vitro* activity of autoantibodies that were identified in our screen and validated by ELISA (**Supplementary Fig. 3a,b**). In a GM-CSF signaling assay based on evaluation of STAT5 phosphorylation in TF-1 cells, we found that purified IgG from a patient with anti-GM-CSF autoantibodies neutralized GM-CSF signaling while purified IgG from uninfected control patients did not **(Fig. 2a)**. Similarly, we assayed chemokine receptor activity using the PRESTO-Tango system^20^ and found that serum-purified IgG from a patient with anti-CXCL7 autoantibodies and a patient with anti-CXCL1 autoantibodies neutralized CXCL7 and CXCL1 signaling on their shared receptor CXCR2, whereas serum-purified IgG from control patients did not **(Fig. 2b,c)**. Thus, these results demonstrate that immune-targeting autoantibodies in COVID-19 patients can directly neutralize the activity of cytokines/chemokines and perturb immune function in affected COVID-19 patients.

**Figure 2:**
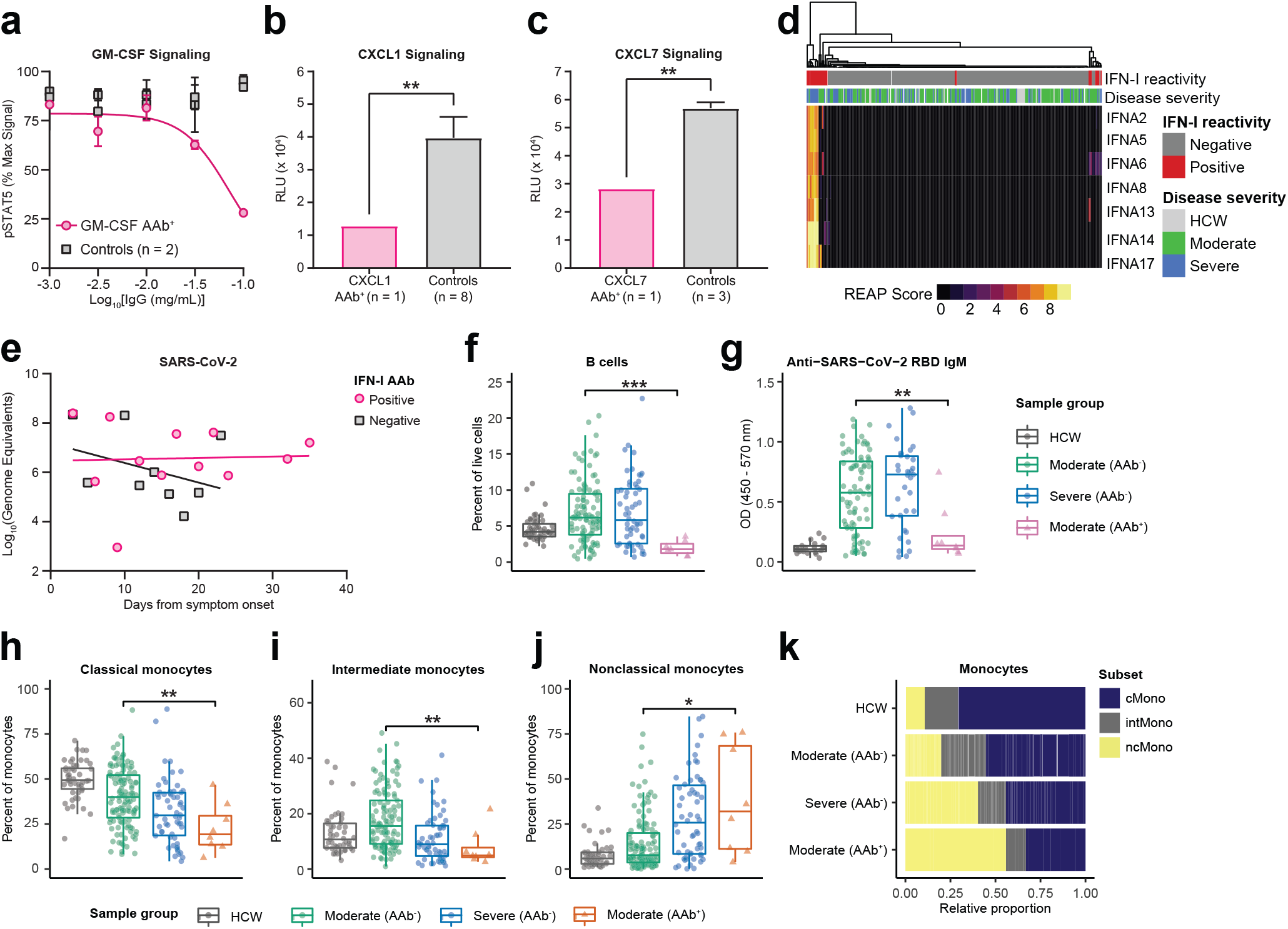
Autoantibodies in COVID-19 patients are functional and correlated with virological and immunological parameters in vivo. **a**, GM-CSF signaling assay based on STAT5 phosphorylation performed in the presence of various concentrations of purified IgG from a COVID-19 patient with GM-CSF autoantibodies and uninfected control plasma samples. Details of percent max signal calculation can be found in methods. Curves were fit using a sigmoidal 4 parameter logistic curve. Results are averages of 2 technical replicates. **b**, CXCL1 and **c**, CXCL7 signaling assay performed in the presence of 0.05 mg/mL purified IgG from a COVID-19 patient with CXCL1 or CXCL7 autoantibodies and uninfected control plasma samples. Results are averages of 3 technical replicates. Significance was determined using a two-sided unpaired t-test (p = 0.0055 in b and 0.0069 in c). **d**, Hierarchically clustered heatmap of IFNα REAP reactivities across all samples. **e**, Longitudinal comparisons of SARS-CoV-2 viral load between patients with and without anti-interferon antibodies. Viral loads were estimated by plotting nasopharyngeal, saliva, or by averaging saliva and nasopharyngeal samples where both were present, in order to generate composite viral loads for each patient. Linear regressions for each group are displayed (solid lines). **f**, Percent B cells among peripheral leukocytes and g, anti-SARS-CoV-2 RBD IgM reactivity as measured by ELISA in samples stratified by COVID-19 disease severity and REAP reactivity (AAb+; REAP score > 2) against B cell displayed proteins (CD38, FcµR, FCRL3). **h-j**, Percentage among total monocytes of classical monocytes (**h**), intermediate monocytes (**i**), and nonclassical monocytes (**j**) in samples stratified by COVID-19 disease severity and REAP reactivity (AAb+; REAP score > 2) against proteins preferentially displayed on classical and intermediate monocytes (CCR2, CCRL2, FFAR4, SYND4, and CPAMD8). Data from **f-j** were presented as boxplots with the first quartile, median, third quartile, and individual data points indicated. **k**, Results from **h-j** represented as horizontal bar charts. Significance was determined using two-sided, Wilcoxon rank-sum test; *P ≤; 0.05, **P ≤; 0.01, ***P ≤; 0.001, ****P ≤; 0.0001. All error bars in this figure represent standard deviation.

To investigate the potential virological effects of cytokine/chemokine targeting autoantibodies, we examined a subset of COVID-19 patients with anti-IFN-I autoantibodies. Confirming a recent report from Bastard et al.^11^, we identified anti-IFN-I autoantibodies in 5.2% of hospitalized COVID-19 patients and additionally found that these autoantibodies were enriched in patients with severe disease **(Fig. 1f, 2d)**. While anti-IFN-I autoantibodies from COVID-19 patients were previously demonstrated to neutralize IFN-α activity *in vitro*, their effect on virological control in infected patients has not been determined^11^. We therefore assessed the functional impact of these autoantibodies in patients by comparing composite viral loads (average of nasopharyngeal and saliva samples) from patients who had anti-IFN-α autoantibodies to those who did not have such antibodies during the course of disease **(Fig. 2e)**. After matching patient groups for comparable average age, sex, and disease severity (average age in anti-IFN-I cohort was 70.25 vs. 71.67 in control; sex distribution was 63% male vs. 67% in control; average disease severity was 3.89 vs. 4.00 in control), patients who had anti-IFN-I antibodies demonstrated impaired virological clearance throughout the course of the study, while patients without anti-IFN-I autoantibodies were able to reduce composite viral loads over time. These results indicate that anti-IFN-I autoantibodies impair the ability to control viral replication in COVID-19 patients.

## Autoantibodies targeting immune cell surface proteins are associated with specific changes in blood leukocyte composition

In addition to their effects on secreted proteins, autoantibodies can also perturb immune responses by binding to targets expressed on the surface of immune cells and triggering opsonization, antibody-dependent cellular cytotoxicity, and/or complement-dependent cytotoxicity^21^. To investigate the effects of autoantibodies against immune cell surface proteins in COVID-19, we looked for associations between these autoantibody reactivities and patient blood leukocyte composition. We found that patients with autoantibodies against antigens expressed on B cells (CD38, FcμR, and FcRL3) had significantly lower frequencies of circulating B-cells when compared to both severity-matched patients without these autoantibodies and control healthcare workers **(Fig. 2f)**. Furthermore, patients with these autoantibodies had significantly lower levels of anti-SARS-CoV-2 RBD IgM when compared to severity-matched patients without these autoantibodies **(Fig. 2g)**. We further examined the patient with anti-CD38 autoantibodies and found that they also exhibited a lower frequency of NK cells, activated CD4^+^ T cells, and activated CD8^+^ T cells, all of which also express CD38 **(Supplementary Fig. 5a-f)**. With respect to monocytes, we identified five autoantibody targets (CCR2, CCRL2, FFAR4, SYND4, and CPAMD8) that were preferentially expressed on classical and intermediate monocytes in a publicly available RNA-seq dataset of human blood leukocytes^22^. We found that patients with these autoantibodies had significantly lower frequencies of classical and intermediate monocytes as well as increased frequencies of nonclassical monocytes when compared to severity-matched patients without these autoantibodies **(Fig. 2h-k)**. Finally, we found one patient with autoantibodies against CD3E (a component of the T cell receptor complex) who had intact B and NK cell compartments but dramatically reduced levels of CD4 T cells, CD8^+^ T cells, and NKT cells in the blood **(Supplementary Fig. 5g-k)**. In aggregate, these data show that autoantibodies that target immune cell surface proteins are associated with depletion of particular immune cell populations and may negatively impact the immune response to SARS-CoV-2.

## Immune-targeting autoantibodies exacerbate disease severity in a mouse model of SARS-CoV-2 infection

To assess the impact of cytokine-targeting autoantibodies in COVID-19 pathogenesis *in vivo*, we used a naturally susceptible mouse model of SARS-CoV-2 infection in which mice transgenically express human ACE2 under the human keratin 18 (*KRT18*) promoter (K18-hACE2)^23^. SARS-CoV-2 infection in K18-hACE2 mice results in robust viral replication and pulmonary inflammation that recapitulate aspects of COVID-19 pathogenesis in human patients^24,25^. Given that anti-IFN-I autoantibodies are enriched in severe COVID-19 patients (**Fig. 2d**), we first administered neutralizing antibodies against the interferon-α/β receptor (IFNAR) in K18-hACE2 mice to examine the impact of antibody-mediated IFN-I blockade *in vivo*. We found that mice pre-treated with anti-IFNAR antibodies were more susceptible to SARS-CoV-2 infection, indicated by increased weight loss (**Fig. 3a**) and reduced survival (**Fig. 3b**). Additionally, in mice treated with anti-IFNAR antibodies, monocyte recruitment, maturation, and differentiation into proinflammatory macrophages in the lung were severely impaired (consistent with the known effect of IFN-I deficiency^26,27^; **Fig. 3c-e**). Furthermore, we found marked increases in the relative frequency (**Fig. 3f**) and absolute number (**Fig. 3g**) of activated lymphoid cells, including CD4^+^ T cells, CD8^+^ T cells, NK cells, and γδ T cells, co-expressing CD44 and CD69 from PBS-treated SARS-CoV-2 infected mice. In contrast, lymphoid cells failed to upregulate activation markers in mice treated with anti-IFNAR antibodies (**Fig. 3f,g**). Collectively, our findings demonstrate that early blockade of IFN-I signaling by antibodies (mimicking the effect of pre-existing anti-IFN-I in patients) interferes with myeloid and lymphoid activation in a murine model of SARS-CoV-2 infection and results in severely exacerbated disease.

**Figure 3:**
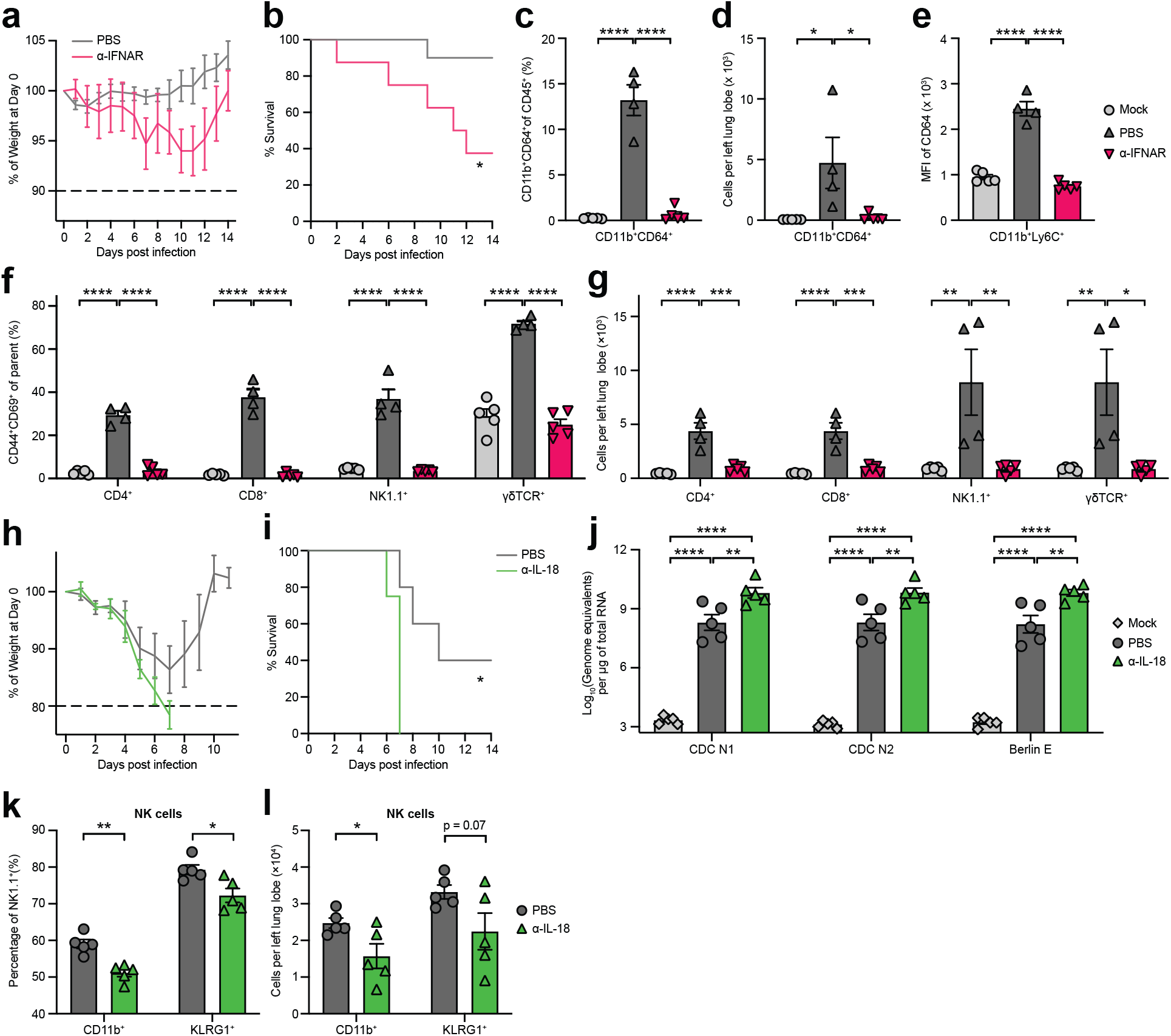
Immune-targeting autoantibodies exacerbate disease in a mouse model of SARS-CoV-2 infection. K18-hACE2 mice were intranasally infected with 103 PFU (**a,b**) or 104 (**c-g, h-l**) PFU SARS-CoV-2. **a**, Body weight of PBS-(n = 10) or αlFNAR-treated (n = 8) K18-hACE2 mice from day 1 to 14 post infection measured as percent of body weight at day 0. **b**, Survival, defined as 10% weight loss, of PBS-(n = 10) or αlFNAR-treated (n = 8) K18-ACE2 from day 1 to 14. **c,d**, Relative frequency (**c**) and abso-lute number (**d**) of lung Ly6C+CD11b+CD64+ macrophages from mock-infected (n = 5), SARS-CoV-2-infected PBS-treated (n = 4), and SARS-CoV-2-infected αlFNAR-treated (n = 5) K18-ACE2 mice. **e**, Expression of CD64 on lung-infiltrating CD11b+Ly6Chigh monocytes from mock-infected (n = 5), SARS-CoV-2-infected PBS-treated (n = 4), and SARS-CoV-2-infected αlFNAR-treated (n = 5) K18-ACE2 mice. **f-g**, Relative frequency (**f**) and absolute number (**g**) of CD44+CD69+ lymphocytes (CD4+ T cells, CD8+ T cells, NK1.1+ cells, and γ^δ^ T cells) measured by flow cytometry. **h**, Body weight of PBS-(n = 5) or αlL-18-treated (n = 4) K18-hACE2 mice from day 1 to 14 post infection measured as percent of body weight at day 0. **i**, Survival, defined as 20% weight loss, of PBS-(n = 5) or alL-18-treated (n = 4) K18-ACE2 from day 1 to 14. **j**, Viral burden from lung tissue homogenates measured 4 days post infection using RT-qPCR primer/probe sets against N or E genes from mock-infected (n = 5), SARS-CoV-2-infected PBS-treated (n = 5), and SARS-CoV-2-infected αlL-18-treated (n = 5) K18-ACE2 mice. **k,l**, Relative frequency (**k**) or absolute number (**l**) of CD11b+ and KLRG1+ NK1.1+ cells in lung tissues from mock-infected (n = 5), SARS-CoV-2-infected PBS-treated (n = 5), and SARS-CoV-2-infected αlL-18-treated (n = 5) K18-ACE2 mice. Significance was determined using two-way ANOVA followed by Sidak correction (**a**), Log-rank (Mantel-Cox) test (**b**), one-way ANOVA followed by Tukey correction (**c-g, j**), and unpaired two-tailed t test (**k,l**); *P ≤ 0.05, **P ≤ 0.01, ***P ≤0.001, ****P ≤ 0.0001. All error bars in this figure represent standard error of the mean.

In addition to anti-IFN-I autoantibodies, we identified autoantibodies against the interleukin-18 (IL-18) pathway, particularly against the IL-18 receptor subunit, IL-18Rβ (*IL18RAP*) (**Supplementary Fig. 3d**). IL-18Rβ serves as a component of the heterodimeric IL-18 receptor (which also contains IL-18Rα), which is a proinflammatory cytokine critical for antiviral NK and CD8^+^ T cell responses^28,29^. To examine the impact of IL-18 pathway disruption in SARS-CoV-2 infection, we administered anti-IL-18 antibodies to K18-hACE2 mice immediately prior to infection. We found that IL-18 blockade greatly enhanced susceptibility to SARS-CoV-2 infection; anti-IL-18-treated mice rapidly lost weight and universally succumbed to the infection, whereas 40% of PBS-treated mice survived (**Fig. 3h,i**). To better understand the mechanisms underlying susceptibility associated with IL-18 blockade, we first measured viral RNA loads in the lung 4 days post-infection. Compared to PBS treatment control, we found that IL-18 blockade resulted in significantly higher viral burden measured by levels of either the N (CDC N1 and CDC N2) or E (Berlin E) gene, suggesting IL-18 is critical for controlling viral replication (**Fig. 3j**). Additionally, given the critical role of IL-18 in inducing the effector properties of NK cells^30^, we measured the expression of various surface markers on NK cells in PBS or anti-IL-18 treated mice. We found that the relative frequency (**Fig. 3k**) and absolute number (**Fig. 3l**) of NK cells expressing CD11b^+^ or KLRG1^+^, which mark the effector subsets with enhanced cytotoxic properties, were significantly reduced in mice treated with anti-IL-18 compared to PBS treated controls. Together, these data demonstrated that antibodies neutralizing the IL-18 pathway resulted in severe impairment in early antiviral control and NK cell responses to SARS-CoV-2 infection in mice.

## Autoantibodies targeting tissue-associated antigens correlate with disease severity and clinical characteristics in COVID-19 patients

In addition to immune-targeting autoantibodies, we also observed a high prevalence of tissue-associated autoantibodies in COVID-19 patients. To understand if these antibodies correlated with particular clinical phenotypes, we manually curated a list of tissue associated antigens with significant differences in REAP signals between uninfected controls and symptomatic patients, and generated a heatmap organized by COVID-19 disease severity (**Fig. 4a**). Broadly, we found a high frequency of autoantibodies directed against the CNS compartment (*e*.*g*., orexin receptor HCRT2R, metabotropic glutamate receptor GRM5, neuronal injury marker NINJ1); against vascular cell types (*e*.*g*., endothelial adhesion molecule PLVAP, regulator of angiogenesis RSPO3); and against connective tissue and extracellular matrix targets (*e*.*g*., suspected regulator of cartilage maintenance OTOR, matrix metalloproteinases MMP7 and MMP9), as well as various other tissue-associated antigens. We next determined whether these tissue-associated antigens had enhanced correlations in severe COVID-19 patients by comparing levels of common clinical laboratory values against REAP scores and generated a difference matrix of Pearson’s r correlation values (moderate correlations subtracted from severe) (**Fig. 4b**). Several tissue groups (*e*.*g*., CNS, Cardiac / Hepatic, Epithelial, Ion Channels) had enhanced correlations with inflammatory clinical markers such as ferritin, C-reactive protein (CRP), and lactate in severe patients relative to moderate COVID-19 cases, high levels of which have been linked to worse COVID-19 disease prognosis^31,32^.

**Figure 4:**
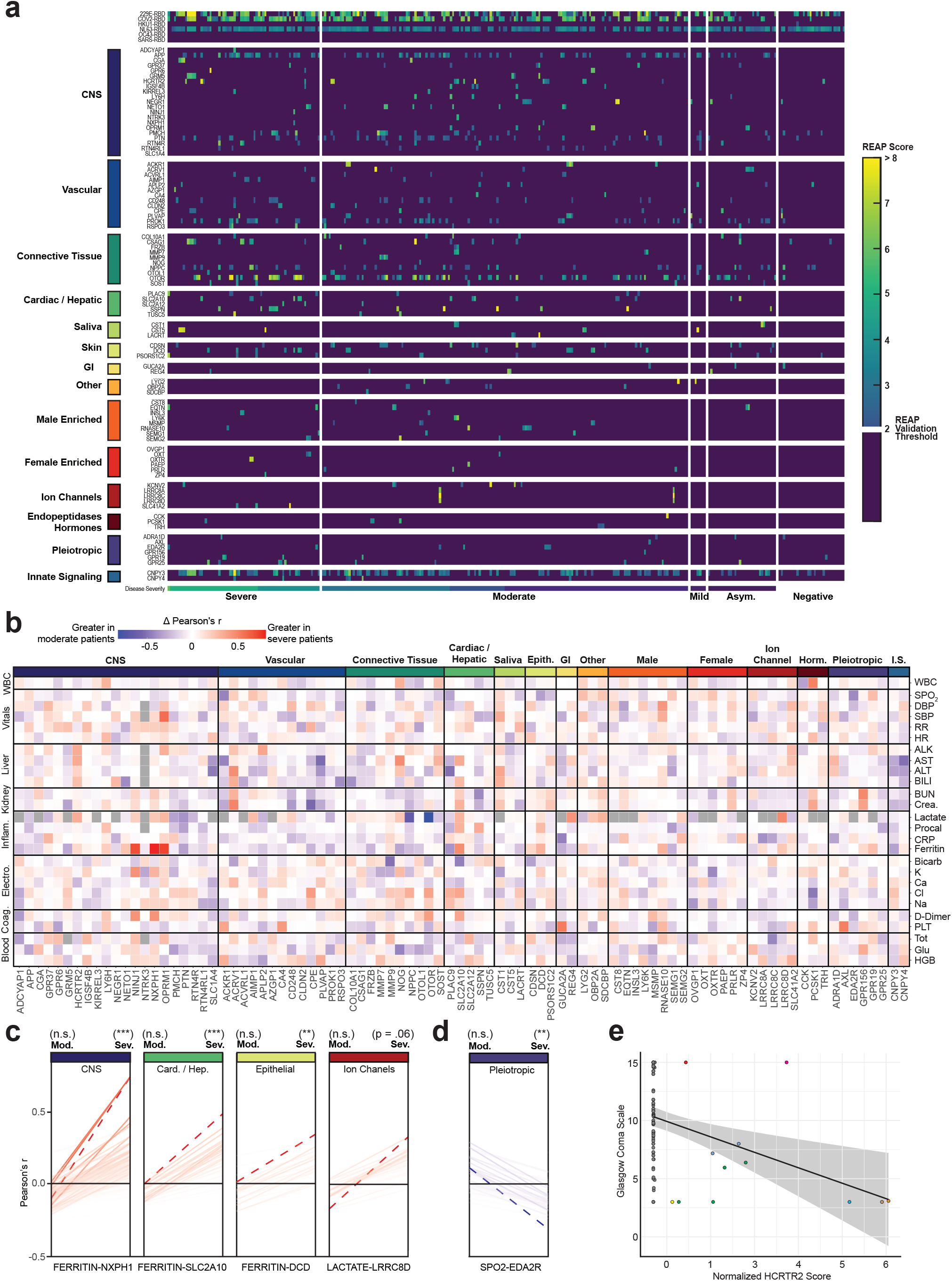
Autoantibodies targeting tissue-associated antigens correlate with disease severity and clinical characteristics in COVID-19 patients. **a**, Heatmap of tissue-associated REAP score stratified by disease severity. Scores below the REAP Validation Threshold of 2.0 were set to 0 to aid interpretation of significant hits. **b**, Difference matrix of Pearson’s r for tissue-associated antigen REAP scores and normalized time-matched clinical laboratory values between severe (n = 93) and moderate (n = 162) COVID-19 patient samples (Δ r = severe r - moderate r). Gray squares indicate r that were unable to be calculated due to missingness of the clinical variable. **c,d**, Change in Pearson’s r for antigen-clinical variable pairs between severe and moderate COVID-19 samples, stratified by positive (red, **c**) and negative (blue, **d**) Δ r. Antigen-clinical variable pairs with greatest Δ r are annotated and indicated with dashed lines (red). Significance of r was determined using two-sided t-tests. **e**, Correlation of normalized HCRTR2 REAP scores with Glasgow Coma Scale scores in severe COVID-19 samples. Blue line shows a linear regression fit. Samples from the same patient were indicated with the same color points. Longitudinal samples from the same patient were included in all analyses in this figure. *P ≤ 0.05, **P ≤ 0.01, ***P ≤ 0.001, ****P ≤ 0.0001.

To systematically identify the greatest changes in autoantibody-clinical variable pairwise correlations, we first stratified antigen-clinical variable pairs into groupings based on positive or negative delta Pearson’s r between moderate and severe COVID-19 patients. We next identified the top-ranked pairings with the greatest changes in correlations between moderate and severe disease states (dashed red line), and found large, significant changes in the correlations for NXPH1, DCD, SLC2A10, and LRRC8D autoreactivity and various inflammatory markers (**Fig. 4c**). We also identified significant changes in the negative correlations for REAP reactivity against the Ectodysplasin A2 receptor (EDA2R), a protein with enhanced expression on type 1 alveolar cells, with oxygen saturation (SpO_2_), suggesting that increasing levels of autoantibodies against this target correlates with decreasing oxygen levels in affected patients (**Fig. 4d**). Given the extent of CNS-specific autoantigens identified in our REAP screen, and recent reports of the potential for SARS-CoV-2 neuroinvasion^33,34^, we additionally examined whether any autoantibodies correlated with individual patient’s Glasgow Coma Scale (GCS) scores. Intriguingly, we found that eight unique COVID-19 patients developed autoantibodies against HCRTR2, an orexin receptor enriched in the hypothalamus. We noted a marked negative correlation between levels of HCRTR2 autoantibodies in these patients and their exceptionally low GCS scores encompassing the time of sample collection (**Fig. 4e**). An additional negative correlation between GRM5 autoreactivity and GCS was found in one of these patients, who eventually succumbed to their infection (**Supplementary Fig. 7**).

## Discussion

The surprising extent of autoantibody reactivities seen in patients with COVID-19 suggests humoral immunopathology is an intrinsic aspect of disease pathogenesis. Screening patient samples with the REAP platform, we identified and validated numerous protein targets involved in a wide range of immunological functions. These autoantibodies had potent functional activities and could be directly correlated with various virological, immunological, and clinical parameters *in vivo* within COVID-19 patient samples. Furthermore, murine surrogates of these autoantibodies led to increased disease severity in a mouse model of SARS-CoV-2 infection. Altogether, these results provide additional evidence that autoantibodies are capable of altering the course of COVID-19 by perturbing the immune response to SARS-CoV-2 and causing direct tissue injury.

Although COVID-19 patients demonstrated high autoreactivity against the exoproteome at a global level, there were essentially no “COVID-19 specific” autoantibodies that distinguished COVID-19 patients from uninfected controls. Similarly, we did not find public responses that could extensively partition patients into specific COVID-19 phenotypes or outcomes. Rather, we observed an extensive constellation of uncommon (*e*.*g*., 5-10% prevalence; anti-IFN-I) and rare (*e*.*g*., <1-5% prevalence; anti-CD38, anti-IL-18Rβ, anti-CD3E) reactivities with large apparent effect sizes. In other words, relatively private reactivities are common in COVID-19, and the aggregate sum of these multifarious responses may explain a significant portion of the clinical variation in patients. It is also intriguing to consider whether the extent of observed autoantibody formation across clinical phenotypes and the absolute breadth of potential targets represents a fundamental defect in tolerance mechanisms as a result of the rapid and exaggerated inflammatory profile attributed to COVID-19. Conversely, patients with pre-existing autoantibodies may be at heightened risk of severe disease due to autoantibody-mediated deficiencies in immune responses during early SARS-CoV-2 infection. Analysis of longitudinal REAP scores within our cohort suggests the existence of both pre-existing autoantibodies, as well as a broad subset of autoantibodies that were induced following infection, indicating that both paradigms may drive the heterogeneity of COVID-19 clinical presentations.

The diversity of autoantibody responses in COVID-19 patients also underscores the importance of high-throughput and unbiased proteome-scale surveys for autoantibody targets. By perturbing biological pathways, autoantibody reactivities are somewhat analogous to genetic mutations^35^ and can uncover unexpected pathways in disease pathophysiology. For instance, beyond validating the biologically-compelling example of anti-IFN-I antibodies in COVID-19, our studies implicated numerous other immune pathways targeted by autoantibodies in COVID-19 that were not previously associated with the disease. In addition to immune-targeting autoantibodies, we also detected antibodies against various tissue-associated antigens. These autoantibodies were correlated with inflammatory clinical markers like ferritin, CRP, and lactate in COVID-19 patients, and these correlations became more extreme with worsening disease progression. Intriguingly, many tissue autoantibodies were present across the diverse physiological compartments frequently implicated during post-COVID syndrome (PCS). Whether the specific autoantibodies identified here play a role in the establishment of PCS, and whether they persist beyond the acute phase of COVID-19, deserves further investigation given the persistent and growing affected patient population.

In summary, our analyses delineated an expansive autoantibody landscape in COVID-19 patients and identified distinct autoantibodies that exerted striking immunological and clinical outcomes. These results implicate previously underappreciated immunological pathways in the etiology of COVID-19 and suggest novel therapeutic paradigms centered around modulating these pathways, as well as attenuating the autoantibodies themselves. Finally, our findings provide a strong rationale for a wider investigation of autoantibodies in infectious disease pathogenesis.

## Materials and Methods

### Ethics statement

This study was approved by Yale Human Research Protection Program Institutional Review Boards (FWA00002571, protocol ID 2000027690). Informed consent was obtained from all enrolled patients and healthcare workers.

### Patients

As previously described^1^ and reproduced here for accessibility, 197 patients admitted to YNHH with COVID-19 between 18 March 2020 and 5 May 2020 were included in this study. No statistical methods were used to predetermine sample size. Nasopharyngeal and saliva samples were collected as described^36^, approximately every four days, for SARS-CoV-2 RT–qPCR analysis where clinically feasible. Paired whole blood for flow cytometry analysis was collected simultaneously in sodium heparin-coated vacutainers and kept on gentle agitation until processing. All blood was processed on the day of collection. Patients were scored for COVID-19 disease severity through review of electronic medical records (EMR) at each longitudinal time point. Scores were assigned by a clinical infectious disease physician according to a custom-developed disease severity scale. Moderate disease status (clinical score 1–3) was defined as: SARS-CoV-2 infection requiring hospitalization without supplementary oxygen (1); infection requiring non-invasive supplementary oxygen (<3 l/min to maintain SpO_2_ >92%) (2); and infection requiring non-invasive supplementary oxygen (>3 l/min to maintain SpO_2_ >92%, or >2 l/min to maintain SpO_2_ >92% and had a high-sensitivity C-reactive protein (CRP) >70) and received tocilizumab). Severe disease status (clinical score 4 or 5) was defined as infection meeting all criteria for clinical score 3 and also requiring admission to the ICU and >6 l/min supplementary oxygen to maintain SpO_2_ >92% (4); or infection requiring invasive mechanical ventilation or extracorporeal membrane oxygenation (ECMO) in addition to glucocorticoid or vasopressor administration (5). Clinical score 6 was assigned for deceased patients. For all patients, days from symptom onset were estimated as follows: (1) highest priority was given to explicit onset dates provided by patients; (2) next highest priority was given to the earliest reported symptom by a patient; and (3) in the absence of direct information regarding symptom onset, we estimated a date through manual assessment of the electronic medical record (EMRs) by an independent clinician. Demographic information was aggregated through a systematic and retrospective review of patient EMRs and was used to construct **Extended Data Table 1**. The clinical data were collected using EPIC EHR and REDCap 9.3.6 software. At the time of sample acquisition and processing, investigators were unaware of the patients’ conditions. Blood acquisition was performed and recorded by a separate team. Information about patients’ conditions was not available until after processing and analysis of raw data by flow cytometry and ELISA. A clinical team, separate from the experimental team, performed chart reviews to determine relevant statistics. Cytokines and FACS analyses were performed blinded. Patients’ clinical information and clinical score coding were revealed only after data collection.

### Clinical Data Acquisition

Clinical data for patients and healthcare workers were extracted from the Yale-New Haven Health computational health platform^37,38^ in the Observational Medical Outcomes Partnership (OMOP) data model. For each research specimen, summary statistics including minimum, mean, median, and maximum values were obtained for relevant clinical measurements, including the Glasgow Coma Scale, within ±1 day from the time of biospecimen collection. Disease severity endpoints, including admission, supplemental oxygen use, and invasive ventilation were validated as previously described^39^.

### Antibodies

Anti-human antibodies used in this study, together with vendors and dilutions, are listed as follows: BB515 anti-hHLA-DR (G46-6) (1:400) (BD Biosciences), BV785 anti-hCD16 (3G8) (1:100) (BioLegend), PE-Cy7 anti-hCD14 (HCD14) (1:300) (BioLegend), BV605 anti-hCD3 (UCHT1) (1:300) (BioLegend), BV711 anti-hCD19 (SJ25C1) (1:300) (BD Biosciences), AlexaFluor 647 anti-hCD1c (L161) (1:150) (BioLegend), Biotin anti-hCD141 (M80), (1:150) (BioLegend), PE-Dazzle594 anti-hCD56 (HCD56) (1:300) (BioLegend), PE anti-hCD304 (12C2) (1:300) (BioLegend), APCFire750 anti-hCD11b (ICRF44) (1:100) (BioLegend), PerCP/Cy5.5 anti-hCD66b (G10F5) (1:200) (BD Biosciences), BV785 anti-hCD4 (SK3) (1:200) (BioLegend), APCFire750 or PE-Cy7 or BV711 anti-hCD8 (SK1) (1:200) (BioLegend), BV421 anti-hCCR7 (G043H7) (1:50) (BioLegend), AlexaFluor 700 anti-hCD45RA (HI100) (1:200) (BD Biosciences), PE anti-hPD1 (EH12.2H7) (1:200) (BioLegend), APC anti-hTIM3 (F38-2E2) (1:50) (BioLegend), BV711 anti-hCD38 (HIT2) (1:200) (BioLegend), BB700 anti-hCXCR5 (RF8B2) (1:50) (BD Biosciences), PECy7 anti-hCD127 (HIL-7R-M21) (1:50) (BioLegend), PE-CF594 anti-hCD25 (BC96) (1:200) (BD Biosciences), BV711 anti-hCD127 (HIL-7R-M21) (1:50) (BD Biosciences), BV785 anti-hCD19 (SJ25C1) (1:300) (BioLegend), BV421 anti-hCD138 (MI15) (1:300) (BioLegend), AlexaFluor700 anti-hCD20 (2H7) (1:200) (BioLegend), AlexaFluor 647 anti-hCD27 (M-T271) (1:350) (BioLegend), PE/Dazzle594 anti-hIgD (IA6-2) (1:400) (BioLegend), PE-Cy7 anti-hCD86 (IT2.2) (1:100) (BioLegend), APC/Fire750 anti-hIgM (MHM-88) (1:250) (BioLegend), BV605 anti-hCD24 (ML5) (1:200) (BioLegend), BV421 anti-hCD10 (HI10a) (1:200) (BioLegend), BV421 anti-CDh15 (SSEA-1) (1:200) (BioLegend), AlexaFluor 700 Streptavidin (1:300) (ThermoFisher), BV605 Streptavidin (1:300) (BioLegend). Anti-mouse antibodies used in this study, together with vendors and dilutions, are listed as follows: FITC anti-mCD11c (N418) (1:400) (BioLegend), PerCP-Cy5.5 or FITC anti-mLy6C (HK1.4) (1:400) (BioLegend), PE or BV605 or BV711 anti-mNK1.1 (PK136) (1:400) (BioLegend), PE-Cy7 anti-mB220 (RA3-6B2) (1:200) (BioLegend), APC anti-mXCR1 (ZET) (1:200) (BioLegend), APC or AlexaFluor 700 or APC-Cy7 anti-mCD4 (RM4-5) (1:400) (BioLegend), APC-Cy7 anti-mLy6G (1A8) (1:400) (BioLegend), BV605 anti-mCD45 (30-F11) (1:400) (BioLegend), BV711 or PerCP-Cy5.5 anti-mCD8a (53-6.7) (1:400) (BioLegend), AlexaFluor 700 or BV785 anti-mCD11b (M1/70) (1:400) (BioLegend), PE anti-mCXCR3 (CXCR3-173) (1:200) (BioLegend), PE-Cy7 anti-mTCRgd (GL3) (1:200) (BioLegend), AlexaFluor 647 anti-mCD19 (6D5) (1:200) (BioLegend), AlexaFluor 700 or BV711 anti-mCD44 (IM7) (1:200) (BioLegend), Pacific Blue anti-mCD69 (H1.2F3) (1:100) (BioLegend), BV605 or APC-Cy7 anti-mCD3 (17A2) (1:200) (BioLegend), BV605 or APC-Cy7 anti-mTCRb (H57-597) (1:200) (BioLegend), BV785 anti-mCD45.2 (104) (1:400) (BioLegend), FITC anti-mKLRG1 (2F1/KLRG1) (1:200) (BioLegend), PE anti-mCD27 (LG.3A10) (1:200) (BioLegend), and Pacific Blue anti-mI-A/I-E (M5/114.15.2) (1:400) (BioLegend).

### Mice

B6.Cg-Tg(K18-ACE2)2Prlmn/J (K18-hACE2) mice were purchased from the Jackson Laboratories and were subsequently bred and housed at Yale University. 5-to 10-week-old mixed sex mice were used throughout the study. All procedures used in this study (sex-matched, age-matched) complied with federal guidelines and the institutional policies of the Yale School of Medicine Animal Care and Use Committee.

### Isolation of plasma

Plasma samples were collected as previously described^1^. Briefly, plasma samples were first isolated from whole blood by centrifugation at 400 *g* for 10 min at room temperature (RT) without brake. The undiluted plasma was then aliquoted and stored at −80 °C for subsequent analysis.

### Isolation of PBMCs

PBMCs were collected as previously described^1^. Briefly, PBMCs were isolated from whole blood by Histopaque (Sigma-Aldrich) density gradient centrifugation. After isolation of plasma, blood was diluted twofold with PBS, layered over Histopaque in a SepMate (StemCell Technologies) tube, underwent centrifugation for 10 min at 1,200 *g*, and further processed according to the manufacturer’s instructions. After PBMC isolation, cells were washed twice with PBS to remove residual Histopaque, treated with ACK Lysing Buffer (ThermoFisher) for 2 min, and then counted. Cell concentration and viability were estimated using standard Trypan blue staining and an automated cell counter (Thermo-Fisher).

### Yeast Induction

All yeast were induced as previously described (Wang et al, manuscript in preparation). In short, one day prior to induction, yeast were expanded in synthetic dextrose medium lacking uracil (SDO -Ura) at 30 °C. The following day, yeast were induced by resuspension at an optical density of 1 in synthetic galactose medium lacking uracil (SGO -Ura) supplemented with 10% SDO - Ura and culturing at 30 °C for approximately 18 hours.

### Yeast Library Construction

The yeast exoproteome library was constructed as previously described (Wang et al, manuscript in preparation). In this study the library was further expanded with the N-terminal extracellular domains of 171 G-protein-coupled receptors (GPCRs) as well as the receptor binding domains (RBDs) of the SARS-CoV-1, SARS-CoV-2, MERS-CoV, HCoV-OC43, HCoV-NL63, and HCoV-229E coronaviruses. Exact sequences of all proteins in the library are provided in **Supplementary Table 1**. Proteins were displayed on yeast as previously described (Wang et al, manuscript in preparation). In short, A two-step PCR process was used to amplify cDNAs for cloning into a barcoded yeast-display vector. cDNA for GPCR N-terminal extracellular domains was derived from the from the PRESTO-Tango plasmid kit, a gift from Bryan Roth (Addgene kit # 1000000068). cDNA for coronavirus RBDs was synthesized by Integrated DNA Technologies. First, cDNAs were amplified with gene-specific primers and PCR products were purified using magnetic PCR purification beads (Avan Bio). Second, the 15bp barcode fragment was constructed by overlap PCR. 4 primers (bc1, bc2, bc3, bc4) were mixed in equimolar ratios and used as a template for a PCR reaction. This PCR product was purified using magnetic PCR purification beads (Avan Bio), reamplified with the first and fourth primer, and then PCR products were run on 2% agarose gels and purified by gel extraction. Purified barcode and gene products were combined with linearized yeast-display vector (pDD003 digested with EcoRI and BamHI) and electroporated into JAR300 yeast using a 96-well electroporator (BTX Harvard Apparatus). Yeast were immediately recovered into 1 mL liquid synthetic dextrose medium lacking uracil (SDO -Ura) in 96-well deep well blocks and grown overnight at 30°C. For coronavirus RBDs, yeast were plated on SDO -Ura agar, single colonies were isolated, and sanger sequencing was used to confirm correct cDNA insertion and identify the associated barcodes. For GPCR N-terminal extracellular domains, these yeast were pooled together with transfected yeast that were used to construct the previously described exoproteome library and a limited dilution of clones were sub-sampled, induced, and stained for FLAG using 1:100 anti-FLAG PE antibody (BioLegend). Yeast were sorted for FLAG display on a Sony SH800Z cell sorter. Barcode-gene pairing for these yeast were performed using a custom Tn5-based sequencing approach as previously described (Wang et al, manuscript in preparation). 4-5 yeast clones for each coronavirus RBD were then spiked into the newly constructed yeast library. All primers used were previously described in Wang et al.

### Rapid Extracellular Antigen Profiling (REAP)

IgG antibody isolation for REAP was performed as previously described (Wang et al, manuscript in preparation). In short, Triton X-100 and RNase A were added to plasma samples at final concentrations of 0.5% and 0.5 mg mL^−1^ respectively and incubated at RT for 30 min before use to reduce risk from any potential virus in plasma. 20 μL protein G magnetic resin (Lytic Solutions) was washed with sterile PBS, resuspended in 75 μL sterile PBS, and added to 25 μL plasma. plasma-resin mixture was incubated overnight at 4 °C with shaking. Resin was washed with sterile PBS, resuspended in 90 µL 100 mM glycine pH 2.7, and incubated for five min at RT. Supernatant was extracted and added to 10 µL sterile 1M Tris pH 8.0. At this point, IgG concentration was measured using a NanoDrop 8000 Spectrophotometer (Thermo Fisher Scientific). This mixture was added to 10^8^ induced empty vector (pDD003) yeast and incubated for 3 hours at 4 °C with shaking. Yeast-IgG mixtures were placed into 96 well 0.45 um filter plates (Thomas Scientific) and yeast-depleted IgG was eluted into sterile 96 well plates by centrifugation at 3000 g for 3 min.

Yeast library selection for REAP was performed as previously described (Wang et al, manuscript in preparation). In short, 400 µL of the induced yeast library was set aside to allow for comparison to post-selection libraries. 10^8^ induced yeast were added to wells of a sterile 96-well v-bottom microtiter plate, resuspended in 100 µL PBE (PBS with 0.5% BSA and 0.5 mM EDTA) containing 10 µg patient-derived antibody, and incubated with shaking for 1 hour at 4 °C. Yeast were washed twice with PBE, resuspended in 100 µL PBE with a 1:100 dilution of biotin anti-human IgG Fc antibody (clone HP6017, BioLegend), and incubated with shaking for 1 hour at 4 °C. Yeast were washed twice with PBE, resuspended in 100 µL PBE with a 1:20 dilution of Streptavidin MicroBeads (Miltenyi Biotec), and incubated with shaking for 30 min at 4 °C. All following steps were carried out at RT. Multi-96 Columns (Miltenyi Biotec) were placed into a MultiMACS M96 Separator (Miltenyi Biotec) in positive selection mode and the columns were equilibrated with 70% ethanol and degassed PBE. Yeast were resuspended in 200 µL degassed PBE and placed into the columns. The columns were washed three times with degassed PBE. To elute the selected yeast, columns were removed from the separator and placed over 96-well deep well plates. 700 µL degassed PBE was added to each well of the column and the column and deep well plate were centrifuged briefly. This process was repeated 3 times. Yeast were recovered in 1 mL SDO -Ura at 30 °C.

DNA was extracted from yeast libraries using Zymoprep-96 Yeast Plasmid Miniprep kits or Zymoprep Yeast Plasmid Miniprep II kits (Zymo Research) according to standard manufacturer protocols. A first round of PCR was used to amplify a DNA sequence containing the protein display barcode on the yeast plasmid. PCR reactions were conducted using 1 µL plasmid DNA, 159_DIF2 and 159_DIR2 primers, and the following PCR settings: 98 °C denaturation, 58 °C annealing, 72 °C extension, 25 rounds of amplification. A second round of PCR was conducted using 1 µL first round PCR product, Nextera i5 and i7 dual-index library primers (Illumina) along with dual-index primers containing custom indices, and the following PCR settings: 98 °C denaturation, 58 °C annealing, 72 °C extension, 25 rounds of amplification. PCR products were pooled and run on a 1% agarose gel. The band corresponding to 257 base pairs was cut out and DNA (NGS library) was extracted using a QIAquick Gel Extraction Kit (Qiagen) according to standard manufacturer protocols. NGS library was sequenced using an Illumina NextSeq 500 and NextSeq 500/550 75 cycle High Output Kit v2.5 with 75 base pair single-end sequencing according to standard manufacturer protocols.

### Recombinant protein production

Recombinant GM-CSF and IL-6 were produced as previously described (Wang et al, manuscript in preparation). In short, sequences encoding the proteins were cloned into a secreted protein expression vector. Expi293 cells (Thermo Fisher Scientific) were transfected and maintained according to manufacturer protocols. 4 days post-transfection, protein was purified from the media by nickel-nitrilotriacetic acid (Ni-NTA) chromatography and desalted into HEPES buffered saline (HBS; 10 mM HEPES, pH 7.5, 150 mM NaCl) + 100 mM sodium chloride. Protein purity was verified by SDS-PAGE.

Recombinant IL-18Rβ was produced as follows. The human IL-18Rβ ectodomain (amino acids 15–356) was cloned into the pACBN_BH3 vector with an N-terminal GP64 signal peptide and a C-terminal AviTag and hexahistidine tag. Plasmids were co-transfected with a transfer vector (Expression Systems, 91002) into Sf9 cells using a baculovirus cotransfection kit (Expression Systems, 91200) per manufacturer’s instructions. P0 virus was harvested after 4-5 days and used for generation of P1/P2 virus stock. For protein production, Hi-5 cells were infected with P2 virus at a previously optimized titer and harvested 3–5 days after infection. Proteins were captured from cell supernatant via Ni-NTA chelating resin and further purified by size exclusion chromatography using an ENrich SEC 650 10 x 300 Column (Bio-Rad) into a final buffer of HEPES buffered saline.

### Autoantibody enzyme-linked immunosorbent assay (ELISA) measurement

200 ng of purchased or independently produced recombinant protein in 100 µL of PBS pH 7.0 was added to 96-well flat-bottom Immulon 2HB plates (Thermo Fisher Scientific) and placed at 4 °C overnight. Plates were washed once with 225 µL ELISA wash buffer (PBS + 0.05% Tween 20) and 150 µL ELISA blocking buffer (PBS + 2% Human Serum Albumin) was added to the well. Plates were incubated for 2 hours at RT. ELISA blocking buffer was removed from the wells and appropriate dilutions of sample plasma in 100 µL ELISA blocking buffer were added to each well. Plates were incubated for 2 hours at RT. Plates were washed 6 times with 225 µL ELISA wash buffer and 1:5000 goat anti-human IgG HRP (Millipore Sigma) or anti-human IgG isotype-specific HRP (Southern Biotech; IgG1: clone HP6001, IgG2: clone 31-7-4, IgG3: clone HP6050, IgG4: clone HP6025) in 100 µL ELISA blocking buffer was added to the wells. Plates were incubated for 1 hour at RT. Plates were washed 6 times with 225 µL ELISA wash buffer. 50 µL TMB substrate (BD Biosciences) was added to the wells and plates were incubated for 20-30 min in the dark at RT. 50 µL 1 M sulfuric acid was added to the wells and absorbance at 450 nm was measured in a Synergy HTX Multi-Mode Microplate Reader (BioTek). Proteins used are as follows: BAMBI (Sino Biological, 10890-H08H-20), C1qB (Sino Biological, 10941-H08B-20), CCL15 (PeproTech, 300-43), CCL16 (PeproTech, 300-44), CD38 (R&D Systems, 2404-AC-010), GM-CSF (produced in-house), CXCL1 (PeproTech, 300-11), CXCL3 (PeproTech, 300-40), CXCL7 (PeproTech, 300-14), FcμR (R&D Systems, 9494-MU-050), IFN-ω (PeproTech, 300-02J), IL-13 (PeproTech, 200-13), IL-1α (RayBiotech, 228-10846-1), IL-6 (produced in-house), TSLP (PeproTech, 300-62), IL-18Rβ (produced in-house).

### SARS-CoV-2 specific antibody ELISA measurement

SARS-CoV-2 specific antibodies were measured as previously described^40^. Briefly, plasma samples were first treated with 0.5% Triton X-100 and 0.5 mg mL^−1^ RNase A at RT for 30 min to inactivate potentially infectious viruses. Meanwhile, recombinant SARS-CoV-2 S1 protein (ACRO Biosystems, S1N-C52H3) or recombinant SARS-CoV-2 RBD protein (ACRO Biosystems, SPD-C82E9) was used to coat 96-well MaxiSorp plates (Thermo Scientific) at a concentration of 2 μg mL^−1^ in PBS in 50 μL per well, followed by overnight incubation at 4 °C. The coating buffer was removed, and plates were incubated for 1 h at RT with 200 μL of blocking solution (PBS with 0.1% Tween-20 and 3% milk powder). Plasma was diluted 1:50 in dilution solution (PBS with 0.1% Tween-20 and 1% milk powder) and 100 μL of diluted serum was added for two hours at RT. Plates were washed three times with PBS-T (PBS with 0.1% Tween-20) and 50 μL of HRP anti-Human IgG Antibody at 1:5000 dilution (GenScript) or anti-Human IgM-Peroxidase Antibody at 1:5000 dilution (Sigma-Aldrich) diluted in dilution solution were added to each well. After 1 h of incubation at RT, plates were washed six times with PBS-T. Plates were developed with 100 μL of TMB Substrate Reagent Set (BD Biosciences 555214) and the reaction was stopped after 12 min by the addition of 100 μL of 2 N sulfuric acid. Plates were then read at a wavelength of 450 nm and 570 nm. The cut-off values for sero-positivity were determined as 0.392, 0.436, and 0.341 for anti-S1-IgG, anti-S1 IgM, and anti-RBD IgG, respectively. 80 and 69 pre-pandemic plasma samples were assayed to establish the negative baselines for the S1 and RBD antigens, respectively. These values were statistically determined with a confidence level of 99%.

### Functional Validation of GM-CSF Autoantibodies

TF-1 cells were starved of recombinant GM-CSF (PreproTech, 300-03) eighteen hours prior to experiments. GM-CSF at 200 pg/mL was incubated with dilutions of purified IgG for 15 min at RT and then used to stimulate TF-1 cells in a 96-well plate (2 × 10^5^ cells per well) in a final volume of 100 µL (final concentration of 100 pg/mL). IgG was purified from plasma using protein G magnetic beads (Lytic Solutions) as previously described (Wang et al, manuscript in preparation). After 15 min of stimulation, cells were fixed in 4% paraformaldehyde for 30 mins, washed with PBS, and permeabilized in 100% methanol on ice for 45 mins. Cells were then washed twice with PBE and stained with PE-conjugated anti-STAT5 pY694 (1:50) (BD Biosciences, 422302) and human TruStain FcX (1:100) (Biolegend, 422302) for 1 hour at RT. Cells were washed with PBE and acquired on a SONY SA3800 flow cytometer. Data were analysed using FlowJo software version 10.6 software (Tree Star). pSTAT5 signal was measured as a function of mean fluorescence intensity (MFI). Percent max signal was calculated by subtracting background MFI and calculating values as a percentage of GM-CSF induced pSTAT5 MFI in the absence of IgG.

### Functional Validation of CXCL1 and CXCL7 Autoantibodies

HTLA cells, a HEK293-derived cell line that stably expresses β-arrestin-TEV and tTA-Luciferase, were seeded in wells of a sterile tissue culture grade flat bottom 96-well plate (35,000 cells/well) in 100 μL DMEM (+ 10% FBS, 1% Penicillin/Streptomycin). 18-24 hr after seeding (approximately 80-90% cell confluence), 200 ng CXCR2-Tango plasmid in 20 μL DMEM and 600 ng Polyethylenimine-Max (Polysciences, 24765-1) in 20 μL DMEM were mixed, incubated at RT for 20 min, and added to each well. 18-24 hr after transfection, medium was replaced with 100 μL DMEM (+ 1% Penicillin/Streptomycin, 10 mM HEPES) containing 10 ng CXCL7 (Peprotech, 300-14) or CXCL1 (PeproTech, 300-46) and 5 ug isolated IgG. IgG was purified from plasma using protein G magnetic beads (Lytic Solutions) as previously described (Wang et al, manuscript in preparation). 18-24 hr after stimulation, supernatant was replaced with 50 μL Bright-Glo solution (Promega) diluted 20-fold with PBS with 20 mM HEPES. The plate was incubated at RT for 20 min in the dark and luminescence was quantified using a Synergy HTX Multi-Mode Microplate Reader (BioTek). HTLA cells were a gift from Noah Palm. CXCR2-Tango was a gift from Bryan Roth (Addgene plasmid # 66260)

### SARS-CoV-2 mouse infections

Before infection, mice were anesthetized using 30% (vol/vol) isoflurane diluted in propylene glycol. 50 µL of SARS-CoV-2 isolate USA-WA1/2020 (NR-52281; BEI Resources) at 2 × 10^4^ or 2 × 10^5^ PFU/mL was delivered intranasally to mice, equivalent of 10^3^ or 10^4^ PFU/mouse, respectively. Following infection, weight loss and survival were monitored daily.

### Viral RNA measurements from human nasopharyngeal samples

RNA concentrations were measured from human nasopharyngeal samples by RT–qPCR as previously described^36^. Briefly, total nucleic acid was extracted from 300 μL of viral transport medium (nasopharyngeal swab) using the MagMAX Viral/Pathogen Nucleic Acid Isolation kit (ThermoFisher) and eluted into 75 μL elution buffer. For SARS-CoV-2 RNA detection, 5 μL of RNA template was tested as previously described^41^, using the US CDC real-time RT–PCR primer/probe sets 2019-nCoV_N1 and 2019-nCoV_N2, as well as the human RNase P (RP) as an extraction control. Virus RNA copies were quantified using a tenfold dilution standard curve of RNA transcripts that we previously generated. If the RNA concentration was lower than the limit of detection (ND) that was determined previously, the value was set to 0 and used for the analyses.

### Viral RNA measurements from mouse lung tissues

Viral RNA from mouse lung tissues was measured as previously described^26^. Briefly, at indicated time points, mice were euthanized with 100% isoflurane. The medial, inferior, and postcaval lobe from the right lung were placed in a Lysing Matrix D tube (MP Biomedicals) with 1 mL of PBS, and homogenized using a table-top homogenizer at medium speed for 2 min. After homogenization, 250 μL of the lung mixture was added to 750 μL Trizol LS (Invitrogen), and RNA was extracted with the RNeasy Mini Kit (Qiagen) according to the manufacturer’s instructions. SARS-CoV-2 RNA levels were quantified with 250 ng of RNA inputs using the Luna Universal Probe Onestep RT-qPCR Kit (New England Biolabs), using real-time RT-PCR primer/probe sets 2019-nCoV_N1 and 2019-nCoV_N2 from the US CDC as well as the primer/probe set E-Sarbeco from the Charité Institute of Virology, Universitätsmedizin Berlin.

### Cytokine and chemokine measurements

Cytokine and chemokine levels from plasma samples were measured as previously described^1^. Briefly, patient serum was isolated as before and aliquots were stored at −80 °C. Sera were shipped to Eve Technologies (Calgary, Alberta, Canada) on dry ice, and levels of cytokines and chemokines were measured using the Human Cytokine Array/Chemokine Array 71-403 Plex Panel (HD71). All samples were measured upon the first thaw.

### Flow cytometry

Flow cytometry on human PBMCs was performed as previously described^1^. Briefly, freshly isolated PBMCs were plated at 1–2 × 10^6^ cells per well in a 96-well U-bottom plate. Cells were resuspended in Live/Dead Fixable Aqua (ThermoFisher) for 20 min at 4 °C. Following a wash, cells were blocked with Human TruStain FcX (BioLegend) for 10 min at RT. Cocktails of desired staining antibodies were added directly to this mixture for 30 min at RT. For secondary stains, cells were first washed and supernatant aspirated; then to each cell pellet a cocktail of secondary markers was added for 30 min at 4 °C. For mouse samples, the left lung was collected at the experimental end point, digested with 1 mg/mL collagenase A (Roche) and 30 µg/mL DNase I (Sigma-Aldrich) in complete RPMI-1640 media for 30 min at 37 °C, and mechanically minced. Digested tissues were then passed through a 70 µm strainer (Fisher Scientific) to single cell suspension and treated with ACK Lysing Buffer (ThermoFisher) to remove red blood cells. Cells were resuspended in Live/Dead Fixable Aqua (ThermoFisher) for 20 min at 4 °C. Following a wash, cells were blocked with anti-mouse CD16/32 antibodies (BioXCell) for 30 min at 4 °C. Cocktails of desired staining antibodies were added directly to this mixture for 30 min at 4 °C. Prior to analysis, human or mouse cells were washed and resuspended in 100 μL 4% PFA for 30–45 min at 4 °C. Following this incubation, cells were washed and prepared for analysis on an Attune NXT (ThermoFisher). Data were analysed using FlowJo software version 10.6 software (Tree Star). The specific sets of markers used to identify each subset of cells are summarized in **Supplementary Fig. 6**.

### Statistical analysis

Specific details of statistical analysis are found in relevant figure legends. Data analysis was performed using R, MATLAB, and GraphPad Prism.

## Data Availability

All non PHI data in the manuscript will be made available upon request.

## Reporting Summary

Further information on research design will be made available in the Nature Research Reporting Summary linked to this article.

## Data Availability

All data generated during this study are available within the paper.

## Acknowledgements

The authors gratefully acknowledge all members of the Ring and Iwasaki labs and also Eric Meffre, Joseph Craft, and Kevin O’Connor for helpful conversation and technical assistance. We thank Melissa Linehan and Huiping Dong for logistical assistance. We also acknowledge H. Patrick Young and Andreas Coppi for their technical assistance with data management. This work was supported by the Mathers Family Foundation (to A.M.R. and A.I.), the Ludwig Family Foundation (to A.M.R. and A.I.), a supplement to the Yale Cancer Center Support Grant 3P30CA016359-40S4 (to A.M.R), the Beatrice Neuwirth Foundation, Yale Schools of Medicine and Public Health and NIAID grant U19 AI08992. IMPACT received support from the Yale COVID-19 Research Resource Fund. A.M.R. is additionally supported by an NIH Director’s Early Independence Award (DP5OD023088) and the Robert T. McCluskey Foundation. T.M. is supported by the Yale Interdisciplinary Immunology Training Program T32AI007019. J.K. is supported by the Yale Medical Scientist Training Program T32GM007205. C.B.F.V. is supported by NWO Rubicon (no. 019.181EN.004).

## Consortia (Yale IMPACT Team)

Abeer Obaid, Adam Moore, Arnau Casanovas-Massana, Alice Lu-Culligan, Allison Nelson, Anderson Brito, Angela Nunez, Anjelica Martin, Annie Watkins, Bertie Geng, Camila D. Odio, Cate Muencker, Chaney Kalinich, Christina Harden, Codruta Todeasa, Cole Jensen, Daniel Kim, David McDonald, Denise Shepard, Edward Courchaine, Elizabeth B. White, Eric Song, Erin Silva, Eriko Kudo, Giuseppe DeIuliis, Harold Rahming, Hong-Jai Park, Irene Matos, Jessica Nouws, Jordan Valdez, Joseph Fauver, Joseph Lim, Kadi-Ann Rose, Kelly Anastasio, Kristina Brower, Laura Glick, Lokesh Sharma, Lorenzo Sewanan, Lynda Knaggs, Maksym Minasyan, Maria Batsu, Mary Petrone, Maxine Kuang, Maura Nakahata, Melissa Campbell, Melissa Linehan, Michael H. Askenase, Michael Simonov, Mikhail Smolgovsky, Nicole Sonnert, Nida Naushad, Pavithra Vijayakumar, Rick Martinello, Rupak Datta, Ryan Handoko, Santos Bermejo, Sarah Prophet, Sean Bickerton, Sofia Velazquez, Tara Alpert, Tyler Rice, William Khoury-Hanold, Xiaohua Peng, Yexin Yang, Yiyun Cao & Yvette Strong

## Author Contributions

E.Y.W., T.M., J.K., Y.D., J.D.H., A.I., and A.M.R. designed experiments. E.Y.W. and Y.D. performed the REAP assay. E.Y.W., Y.D., J.D.H., F.L., E.S.P., and S.F. performed biochemical and functional validations. T.M., B.I., and E.S. performed mouse experiments. C.L., P.W., J.K., J.S., T.M., and J.E.O. defined parameters for flow cytometry experiments, collected and processed patient PBMC samples. A.L.W., C.B.F.V., I.M.O., C.C.K., M.E.P., and A.E.W. performed the virus RNA concentration assays. N.D.G. supervised the virus RNA concentration assays. B.I. and J.K. collected epidemiological and clinical data. C.D.C., S.F., A.I.K., M.C., and J.F. assisted in designing, recruiting, and following in-patient and HCW cohorts. W.L.S. supervised clinical data management. E.Y.W., T.M., J.K., N.S.Z., Y.D., J.D.H., A.I., and A.M.R. analyzed data. E.Y.W., T.M., J.K., A.I., and A.M.R. wrote the paper. A.M.R. and A.I. supervised the research. Authors from the Yale IMPACT Research Team contributed to collection and storage of patient samples, as well as the collection of the patients’ epidemiological and clinical data.

## Corresponding Authors

Correspondence to Akiko Iwasaki or Aaron M. Ring.

---

**Extended Data Table 1.**
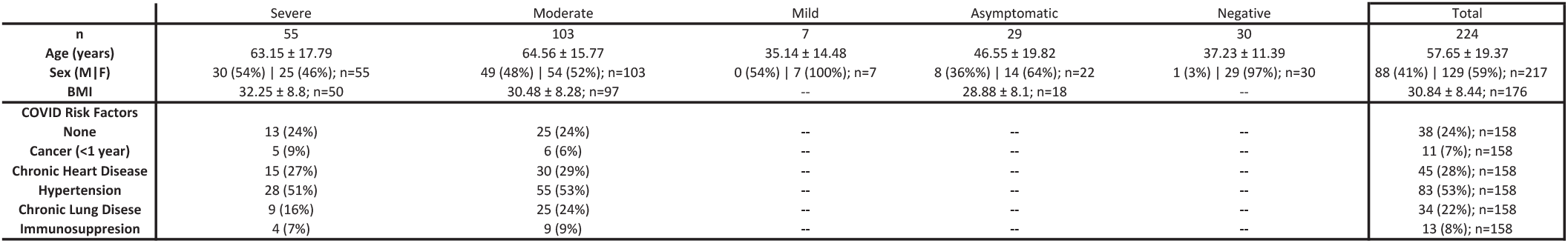

**Supplementary Figure 1.**
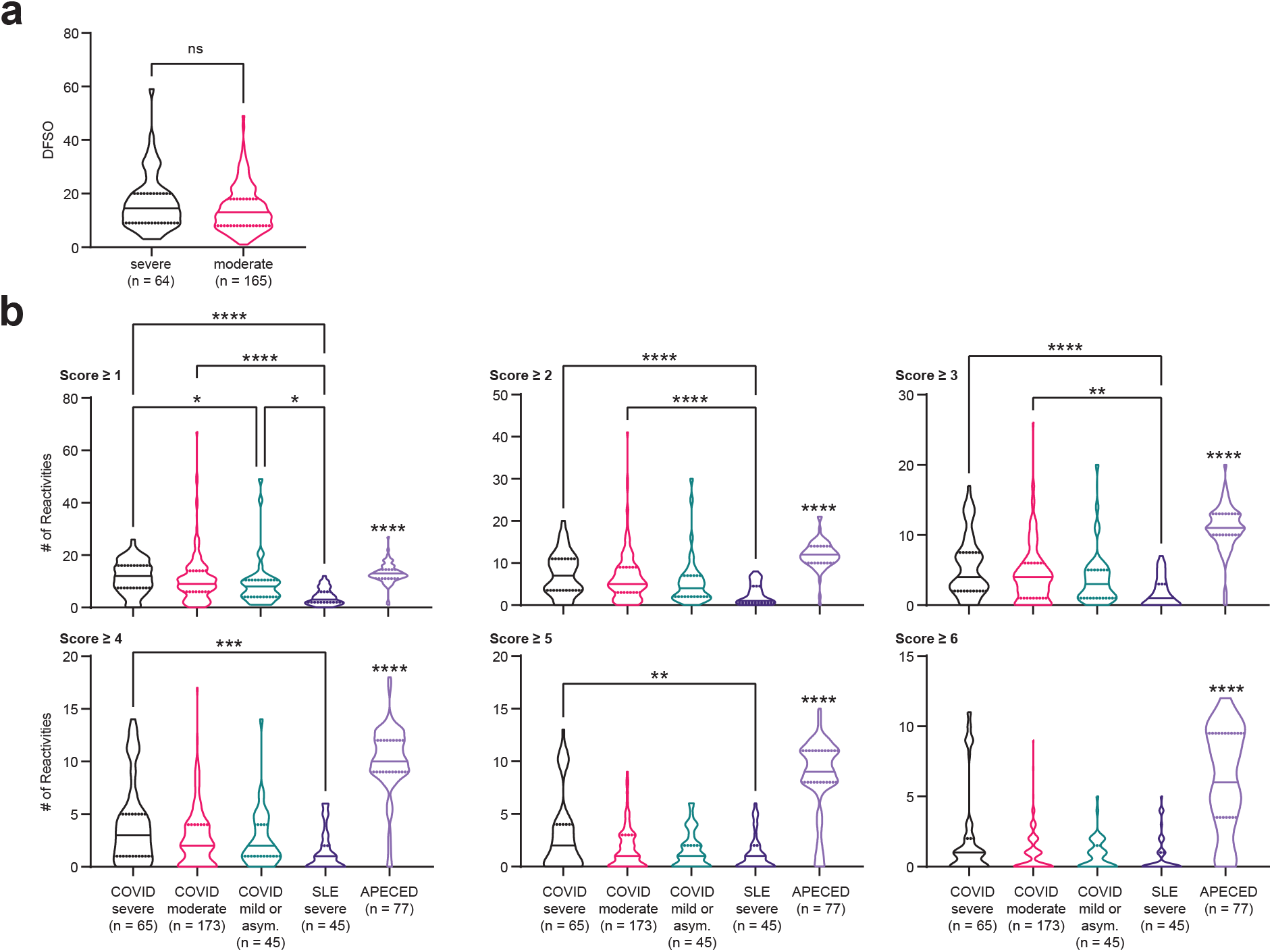
**a**, Violin plots of days from symptom onset (DFSO) in severe and moderate COVID-19 samples. DFSO data was not available for a limited number of samples from each group. Significance was determined using a two-sided Mann-Whitney U test. **b**, Violin plots of the number of reactivities in severe COVID-19, moderate COVID-19, mild or asymptomatic COVID-19, severe SLE, and APECED patient samples at different score cutoffs. SLE and APECED patients were screened as previously described (Wang et al, manuscript in preparation). Due to the smaller size of the yeast exoproteome library used to screen the SLE and APECED samples, reactivities in the COVID-19 cohort against proteins that were not in the previously described yeast exoproteome library were removed from these analyses. Significance was determined using a Kruskal-Wallis test followed by a Dunn’s test. Significance indicators above the APECED group represent significance in comparison to all other groups. In all violin plots in this figure, solid lines represent the median and dotted lines represent the first or third quartile. **P ≤ 0.01, ***P ≤ 0.001, ****P ≤ 0.0001.

**Supplementary Figure 2.**
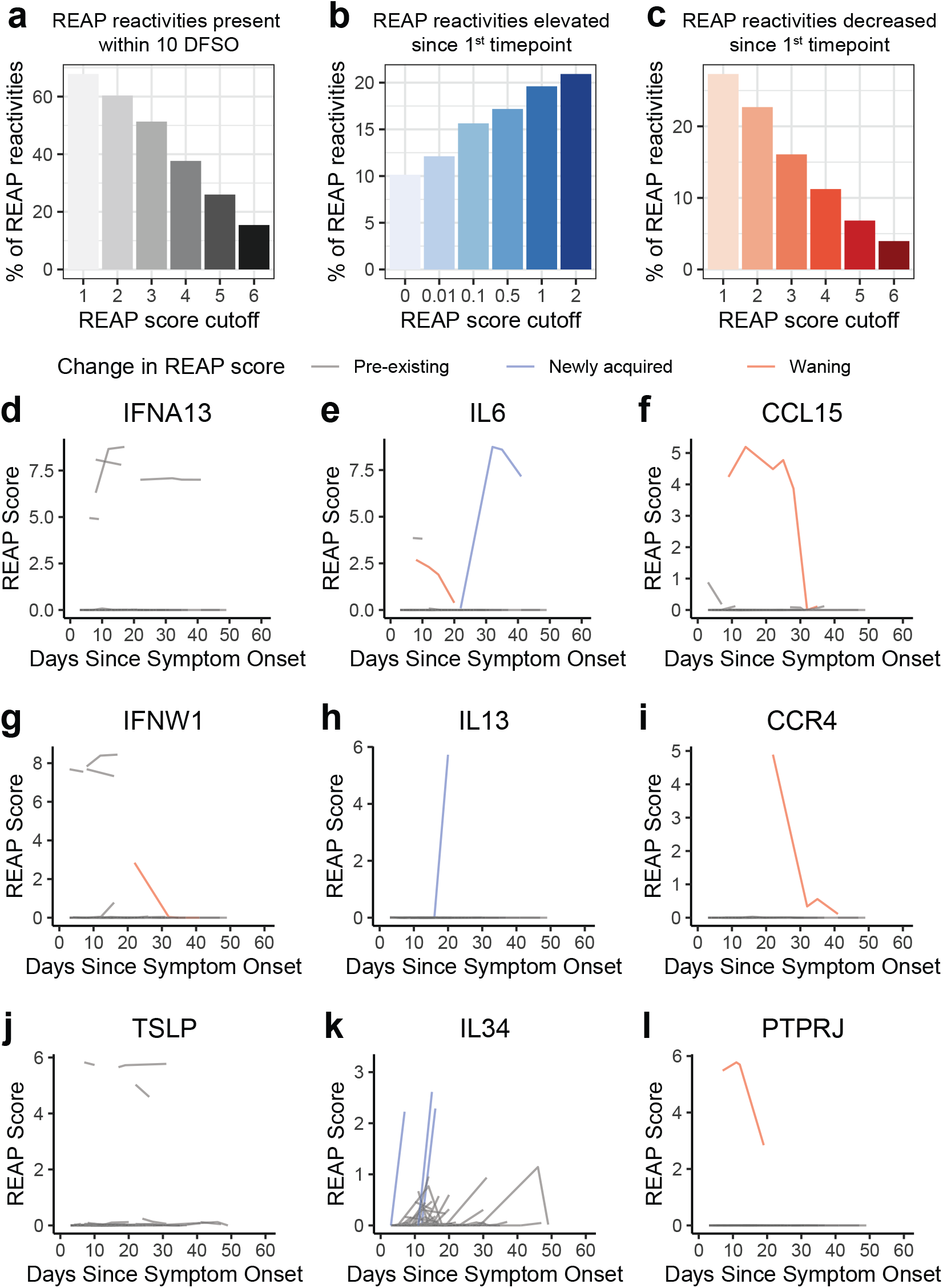
**a**, Percentage of reactivities (REAP score greater than score cutoff) in COVID-19 patients present within 10 days from symptom onset at various score cutoffs. **b**, Percentage of reactivities in COVID-19 patients that had a REAP score less than the score cutoff (using various score cutoffs) at the first time point sampled and an increase in REAP score of at least 1 at the last time point. **c**, Percentage of reactivities in COVID-19 patients that had a REAP score greater than the score cutoff (using various score cutoffs) at the first time point sampled and a decrease in REAP score of at least 1 at the last time point. **d-l**, Plots of longitudinal changes in REAP score for autoreactivities against IFNA13 (**d**), IL6 (**e**), CCL15 (**f**), IFNW1 (**g**), IL13 (**h**), CCR4 (**i**), TSLP (**j**), IL34 (**k**), and PTPRJ (**l**) in COVID-19 patients.

**Supplementary Figure 3.**
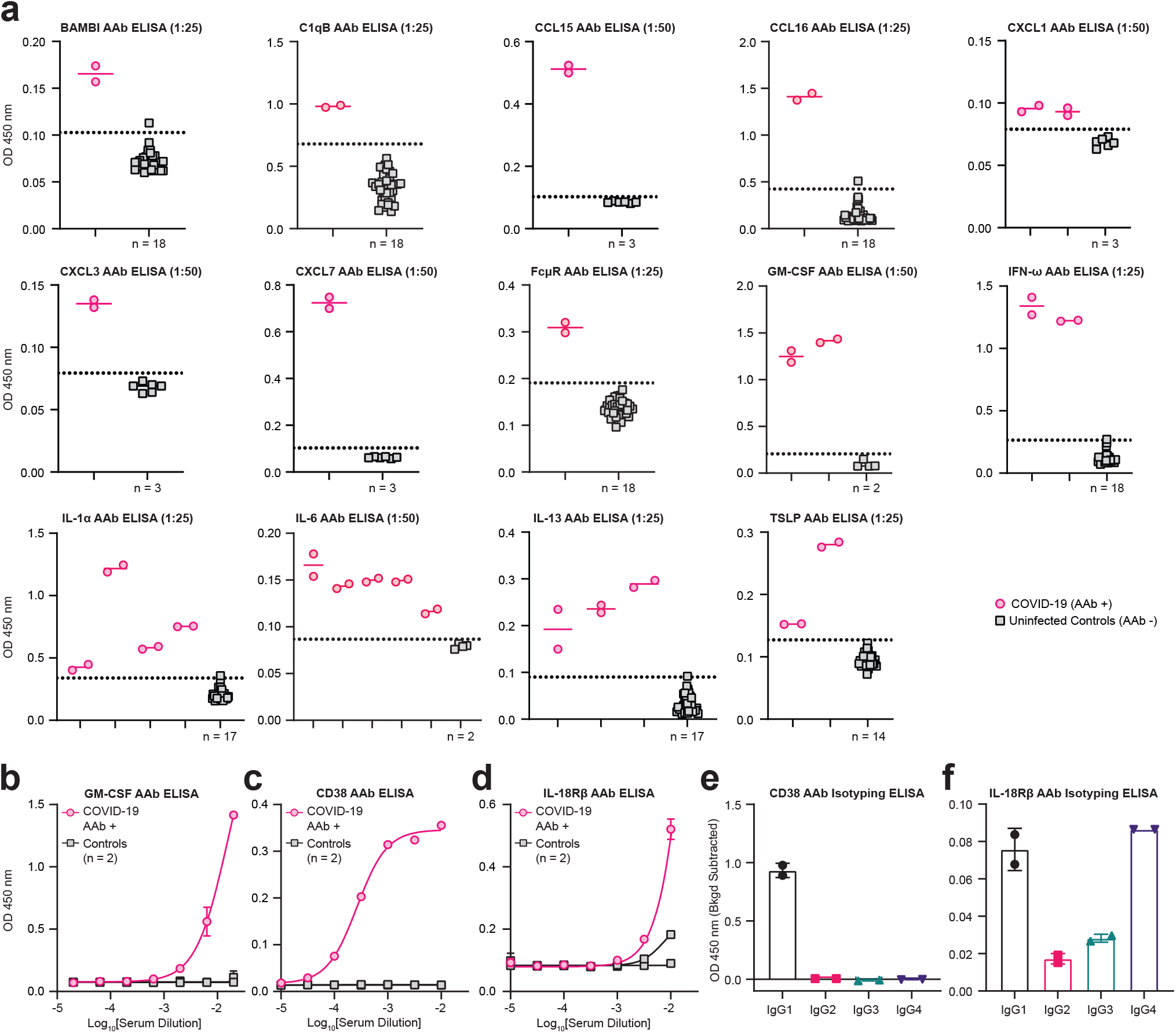
**a**, Single point pan-IgG autoantibody ELISAs conducted 1:25 or 1:50 plasma dilution (indicated in graph titles). Dotted line represents the healthy donor average plus 3 standard deviations. The number of unique controls used in each ELISA is indicated below the control column in each graph. Technical replicates are depicted as distinct points on graphs. **b**, GM-CSF, **c**, CD38, and **d**, IL-1SR β pan-IgG autoantibody ELISAs conducted with serial dilutions of COVID-19 patient or uninfected control plasma. Technical replicates were performed for all dilutions and samples. Error bars represent standard deviation. Results are averages of 2 technical replicates. Curves were fit using a sigmoidal 4 parameter logistic curve. **e**, CD38 and **f**, IL-1SRβ IgG subclass specific ELISAs conducted with 1:100 plasma dilution. Technical replicates are depicted as distinct points on graphs. Background optical density (OD) values were subtracted to normalize for varying background levels of the subclass specific secondary antibodies. All error bars in this figure represent standard deviation.

**Supplementary Figure 4.**
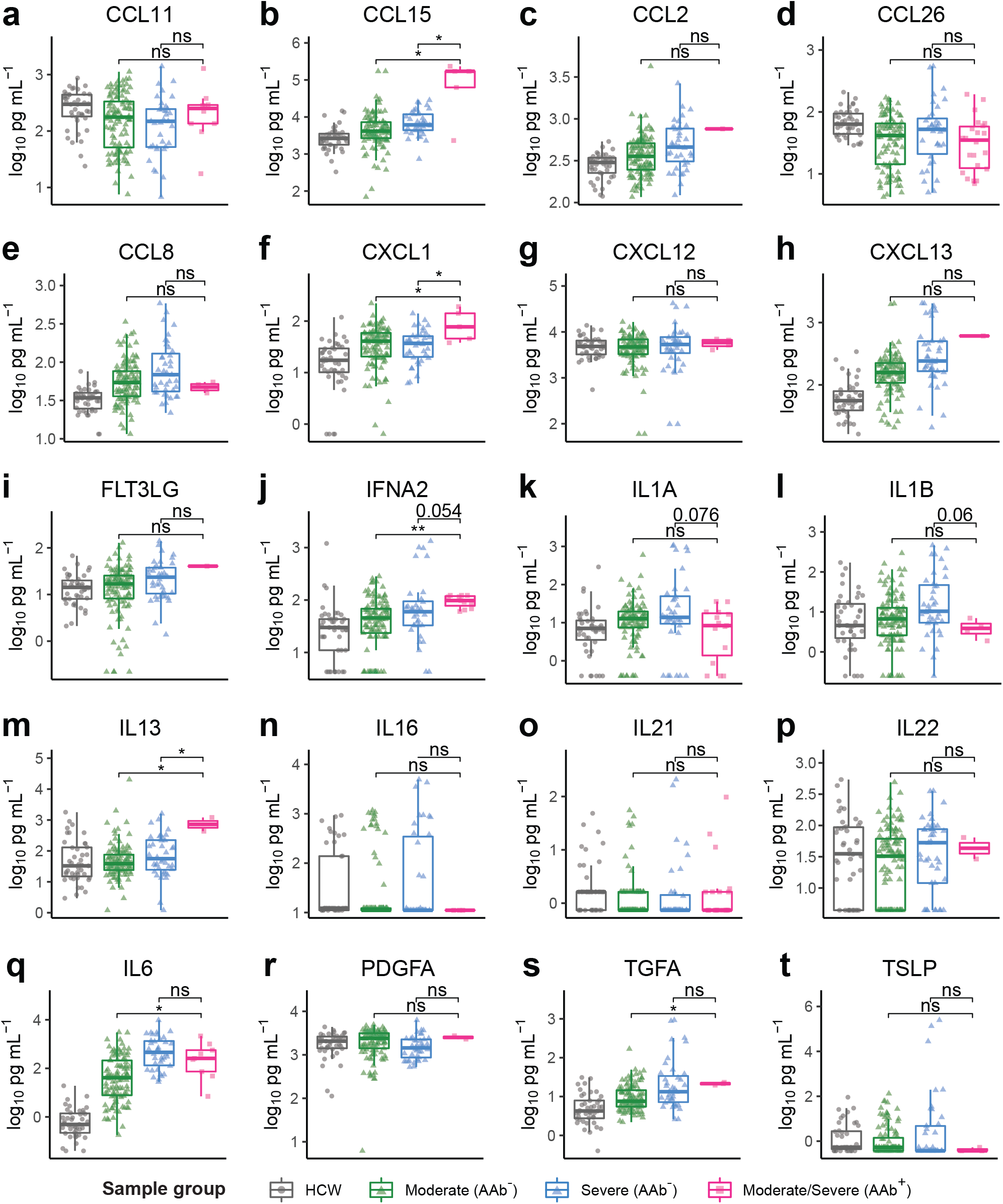
**a-t**, Concentration of plasma CCL11 (**a**), CCL15 (**b**), CCL2 (**c**), CCL26 (**d**), CCL8 (**e**), CXCL1 (**f**), CXCL12 (**g**), CXCL13 (**h**), FLT3LG (**i**), IFNA2 (**j**), IL1A (**k**), IL1B (**l**), IL13 (**m**), IL16 (**n**) and IL21 (**o**), IL22 (**p**), IL6 (**q**), PDGFA (**r**), TGFA (**s**), and TSLP (**t**) measured by a Luminex assay in samples stratified by COVID-19 disease severity and REAP reactivity (AAb+; REAP score >= 2) against the corresponding cytokine. Significance was determined using two-sided, Wilcoxon rank-sum test *P ≤ 0.05, **P ≤ 0.01, ***P ≤ 0.001, ****P ≤ 0.0001. All error bars in this figure represent standard deviation.

**Supplementary Figure 5.**
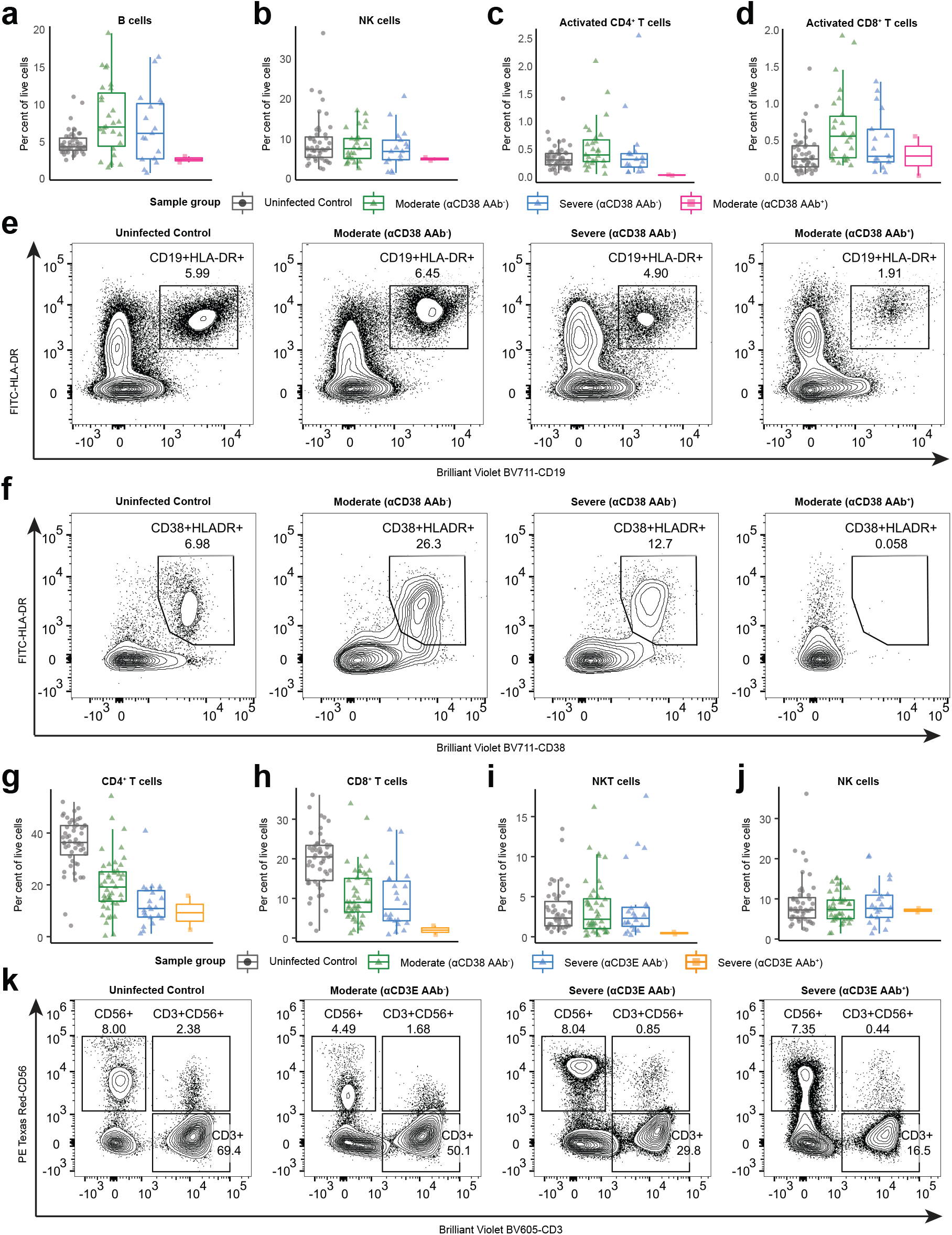
**a-d**, Percent B cells (**a**), NK cells (**b**), Activated CD4+ T cells (**c**), activated CDS+ T cells (**d**) among peripheral leukocytes in samples stratified by COVID-19 disease severity and REAP reactivity (score >= 2) to CD3S. **e**, Representative flow plot of B cells (CD19+HLA-DR+) for **a. f**, Representative flow plot of activated CDS+ T cells cells (CD19+HLA-DR+) for **d. g-j**, Percent CD4+ T cells (**g**), CDS+ T cells (**h**), NKT cells (**i**), and NK cells (**j**) among peripheral leukocytes in samples stratified by COVID-19 disease severity and REAP reactivity (score >= 2) to CD3E. **k**, Representative flow plot of T cells (CD3+), NK cells (CD56+), and NKT cells (CD3+CD56+) for **g-j**.

**Supplementary Figure 6.**
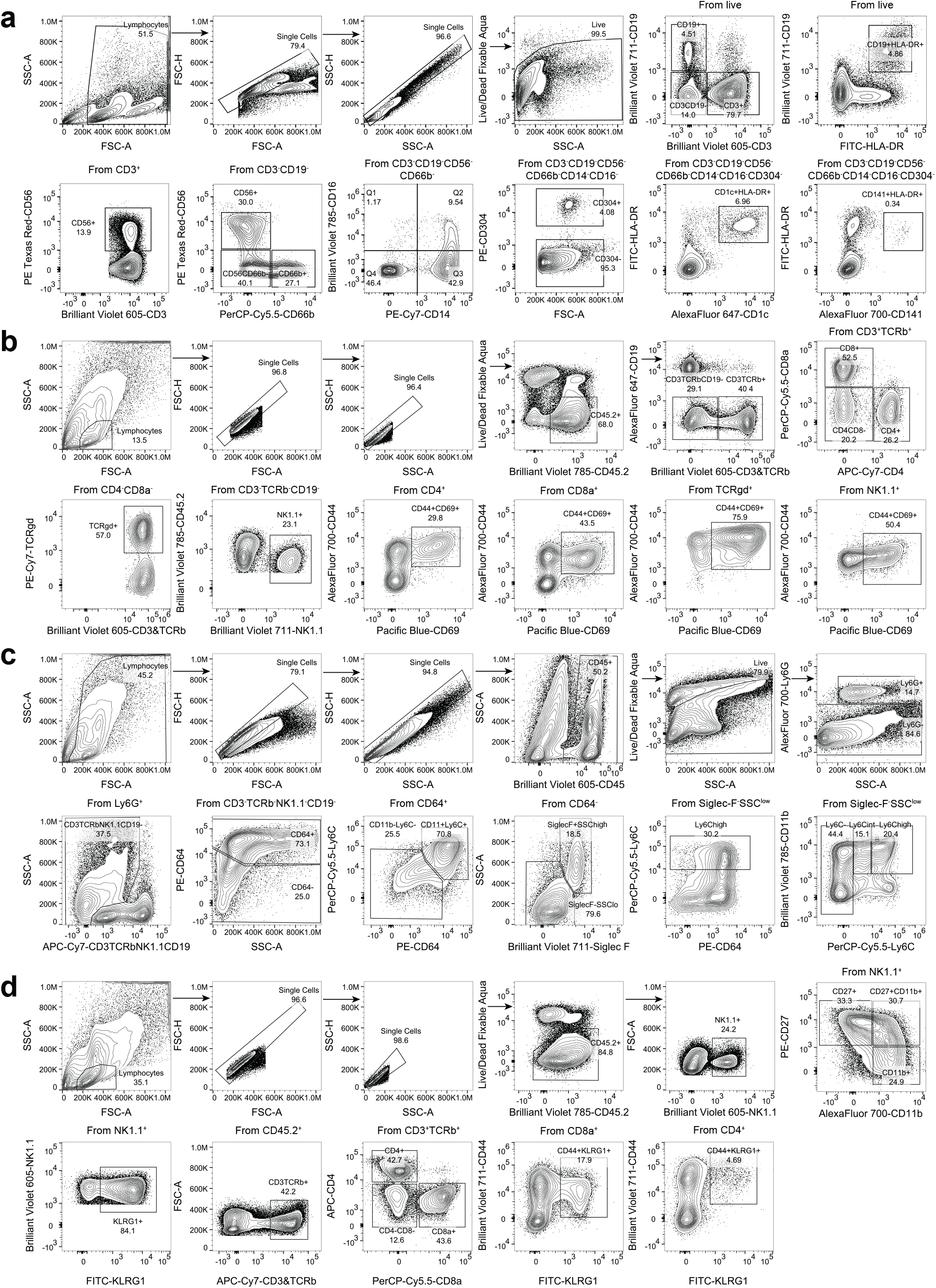
**a**, Gating strategy to identify B cells in Fig. 2f, monocytes in **Fig. 2h-k**, T cells, NKT cells, and NK cells in **Supplementary Fig. 5**. in human PBMCs. **b**, Gating strategy to identify CD11b+Ly6Chigh monocytes and Ly6C+CD11b+CD64+ macrophages in mouse lung tissues described in **Fig. 3c-e. c**, Gating strategy to identify CD44+CD69+ lymphocytes in mouse lung tissues described in **Fig. 3f-g. d**, Gating strategy to identify KLRG1+ and CD11b+ NK cells described in **Fig. 3k-l**.

**Supplementary Figure 7.**
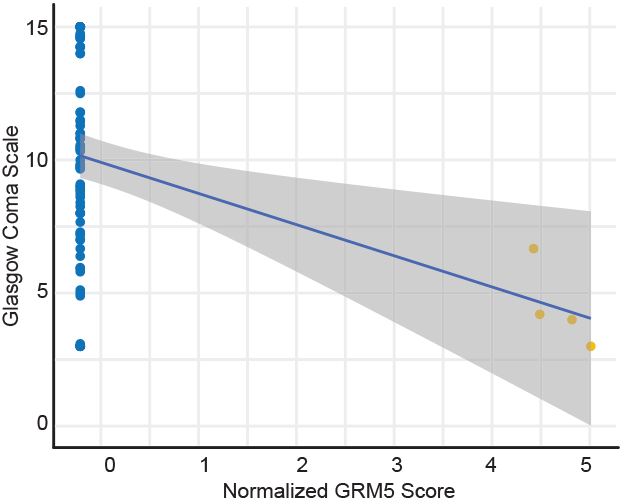
Correlation of normalized GRM5 REAP scores with Glasgow Coma Scale scores in severe COVID-19 samples. Blue line shows a linear regression fit. Samples from the same patient were indicated with the same color points.

