## Supplementary Figures for "Diverse Functional Autoantibodies in Patients with COVID-19"

### Extended Data Table 1

| | Severe | Moderate | Mild | Asymptomatic | Negative | Total |
|---|---|---|---|---|---|---|
| **n** | 55 | 103 | 7 | 29 | 30 | 224 |
| **Age (years)** | 63.15 ± 17.79 | 64.56 ± 15.77 | 35.14 ± 14.48 | 46.55 ± 19.82 | 37.23 ± 11.39 | 57.65 ± 19.37 |
| **Sex (M\|F)** | 30 (54%) \| 25 (46%); n=55 | 49 (48%) \| 54 (52%); n=103 | 0 (54%) \| 7 (100%); n=7 | 8 (36%%) \| 14 (64%); n=22 | 1 (3%) \| 29 (97%); n=30 | 88 (41%) \| 129 (59%); n=217 |
| **BMI** | 32.25 ± 8.8; n=50 | 30.48 ± 8.28; n=97 | -- | 28.88 ± 8.1; n=18 | -- | 30.84 ± 8.44; n=176 |
| **COVID Risk Factors** | | | | | | |
| **None** | 13 (24%) | 25 (24%) | -- | -- | -- | 38 (24%); n=158 |
| **Cancer (<1 year)** | 5 (9%) | 6 (6%) | -- | -- | -- | 11 (7%); n=158 |
| **Chronic Heart Disease** | 15 (27%) | 30 (29%) | -- | -- | -- | 45 (28%); n=158 |
| **Hypertension** | 28 (51%) | 55 (53%) | -- | -- | -- | 83 (53%); n=158 |
| **Chronic Lung Disese** | 9 (16%) | 25 (24%) | -- | -- | -- | 34 (22%); n=158 |
| **Immunosuppresion** | 4 (7%) | 9 (9%) | -- | -- | -- | 13 (8%); n=158 |

### Supplementary Figure 1

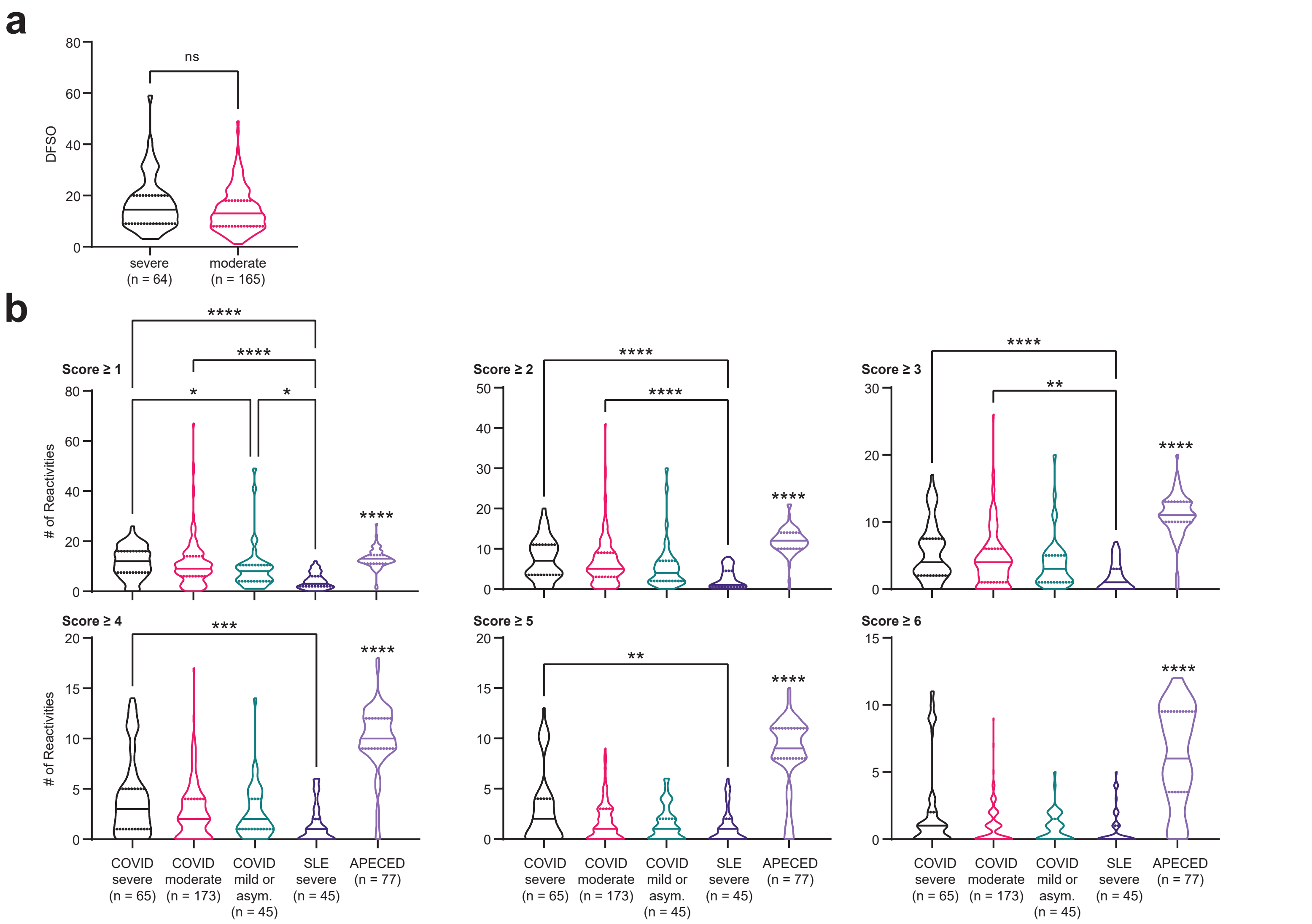


**Supplementary Figure 1. a,** Violin plots of days from symptom onset (DFSO) in severe and moderate COVID-19 samples. DFSO data was not available for a limited number of samples from each group. Significance was determined using a two-sided Mann-Whitney U test. **b,** Violin plots of the number of reactivities in severe COVID-19, moderate COVID-19, mild or asymptomatic COVID-19, severe SLE, and APECED patient samples at different score cutoffs. SLE and APECED patients were screened as previously described (Wang et al, manuscript in preparation). Due to the smaller size of the yeast exoproteome library used to screen the SLE and APECED samples, reactivities in the COVID-19 cohort against proteins that were not in the previously described yeast exoproteome library were removed from these analyses. Significance was determined using a Kruskal-Wallis test followed by a Dunn's test. Significance indicators above the APECED group represent significance in comparison to all other groups. In all violin plots in this figure, solid lines represent the median and dotted lines represent the first or third quartile. $**P \leq 0.01$, $***P \leq 0.001$, $****P \leq 0.0001$.

### Supplementary Figure 2

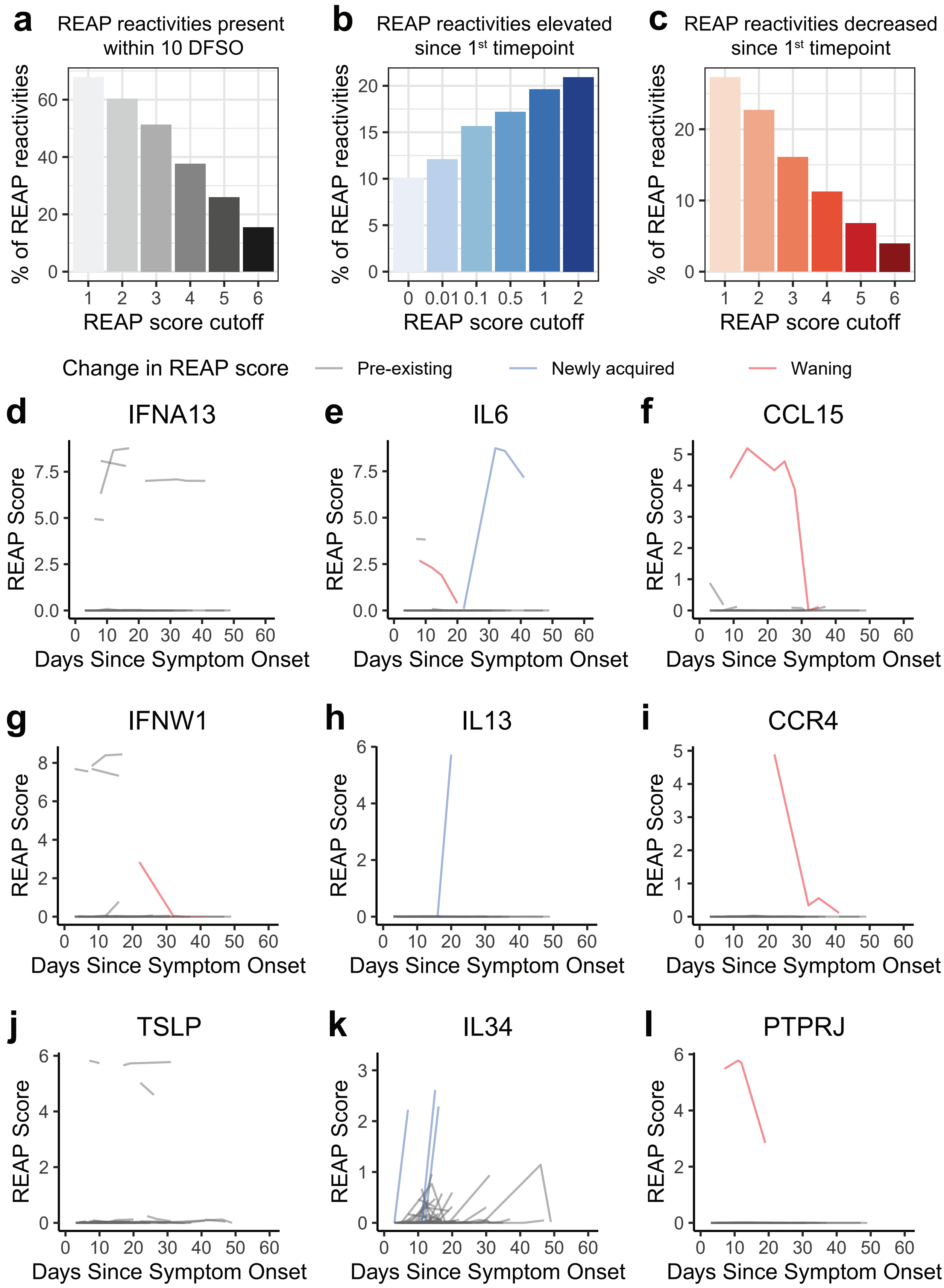


**Supplementary Figure 2. a,** Percentage of reactivities (REAP score greater than score cutoff) in COVID-19 patients present within 10 days from symptom onset at various score cutoffs. **b,** Percentage of reactivities in COVID-19 patients that had a REAP score less than the score cutoff (using various score cutoffs) at the first time point sampled and an increase in REAP score of at least 1 at the last time point. **c,** Percentage of reactivities in COVID-19 patients that had a REAP score greater than the score cutoff (using various score cutoffs) at the first time point sampled and a decrease in REAP score of at least 1 at the last time point. **d-l,** Plots of longitudinal changes in REAP score for autoreactivities against IFNA13 (**d**), IL6 (**e**), CCL15 (**f**), IFNW1 (**g**), IL13 (**h**), CCR4 (**i**), TSLP (**j**), IL34 (**k**), and PTPRJ (**l**) in COVID-19 patients.

### Supplementary Figure 3

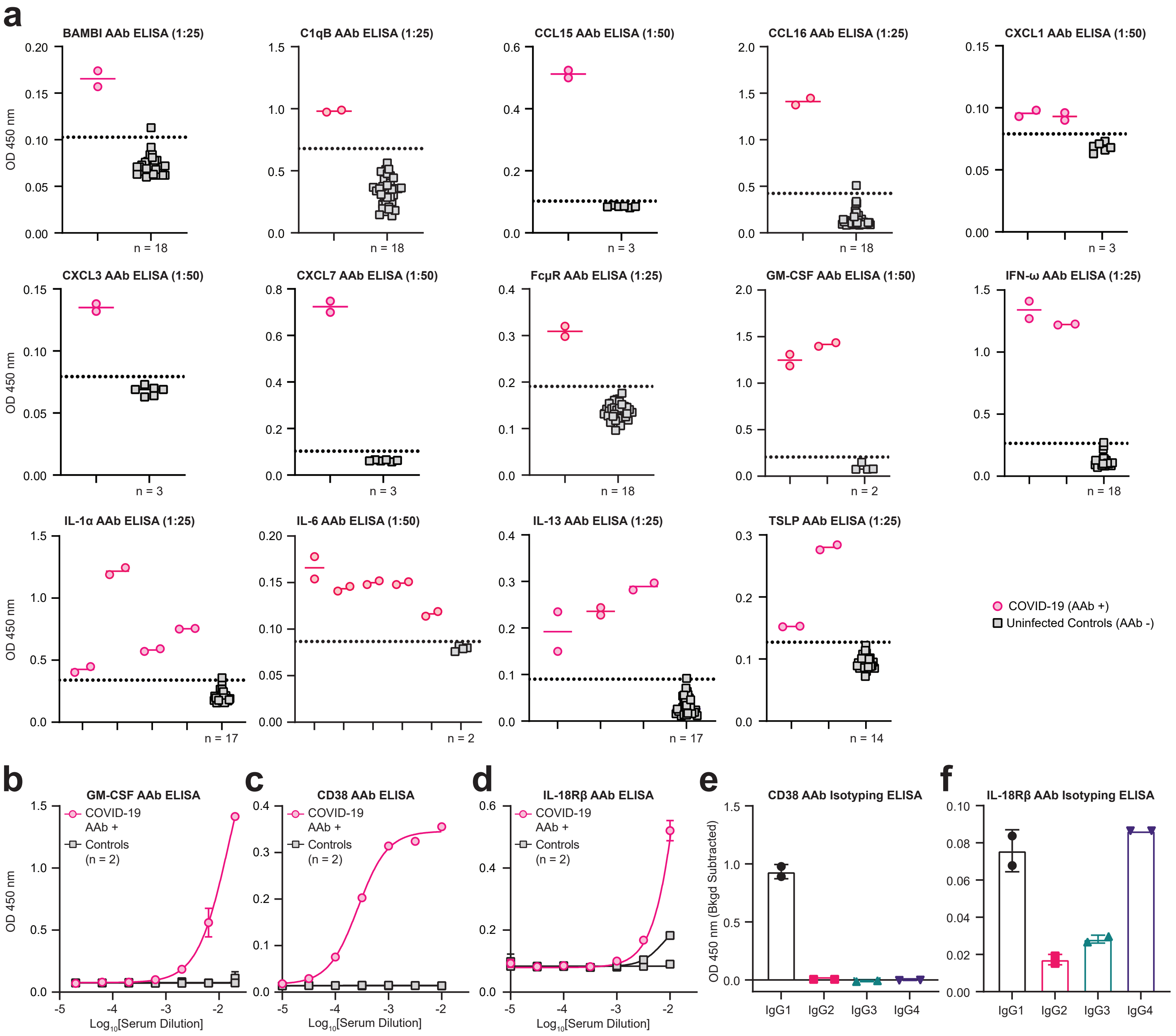


**Supplementary Figure 3. a,** Single point pan-IgG autoantibody ELISAs conducted 1:25 or 1:50 plasma dilution (indicated in graph titles). Dotted line represents the healthy donor average plus 3 standard deviations. The number of unique controls used in each ELISA is indicated below the control column in each graph. Technical replicates are depicted as distinct points on graphs. **b,** GM-CSF, **c,** CD38, and **d,** IL-18Rβ pan-IgG autoantibody ELISAs conducted with serial dilutions of COVID-19 patient or uninfected control plasma. Technical replicates were performed for all dilutions and samples. Error bars represent standard deviation. Results are averages of 2 technical replicates. Curves were fit using a sigmoidal 4 parameter logistic curve. **e,** CD38 and **f,** IL-18Rβ IgG subclass specific ELISAs conducted with 1:100 plasma dilution. Technical replicates are depicted as distinct points on graphs. Background optical density (OD) values were subtracted to normalize for varying background levels of the subclass specific secondary antibodies. All error bars in this figure represent standard deviation.

### Supplementary Figure 4

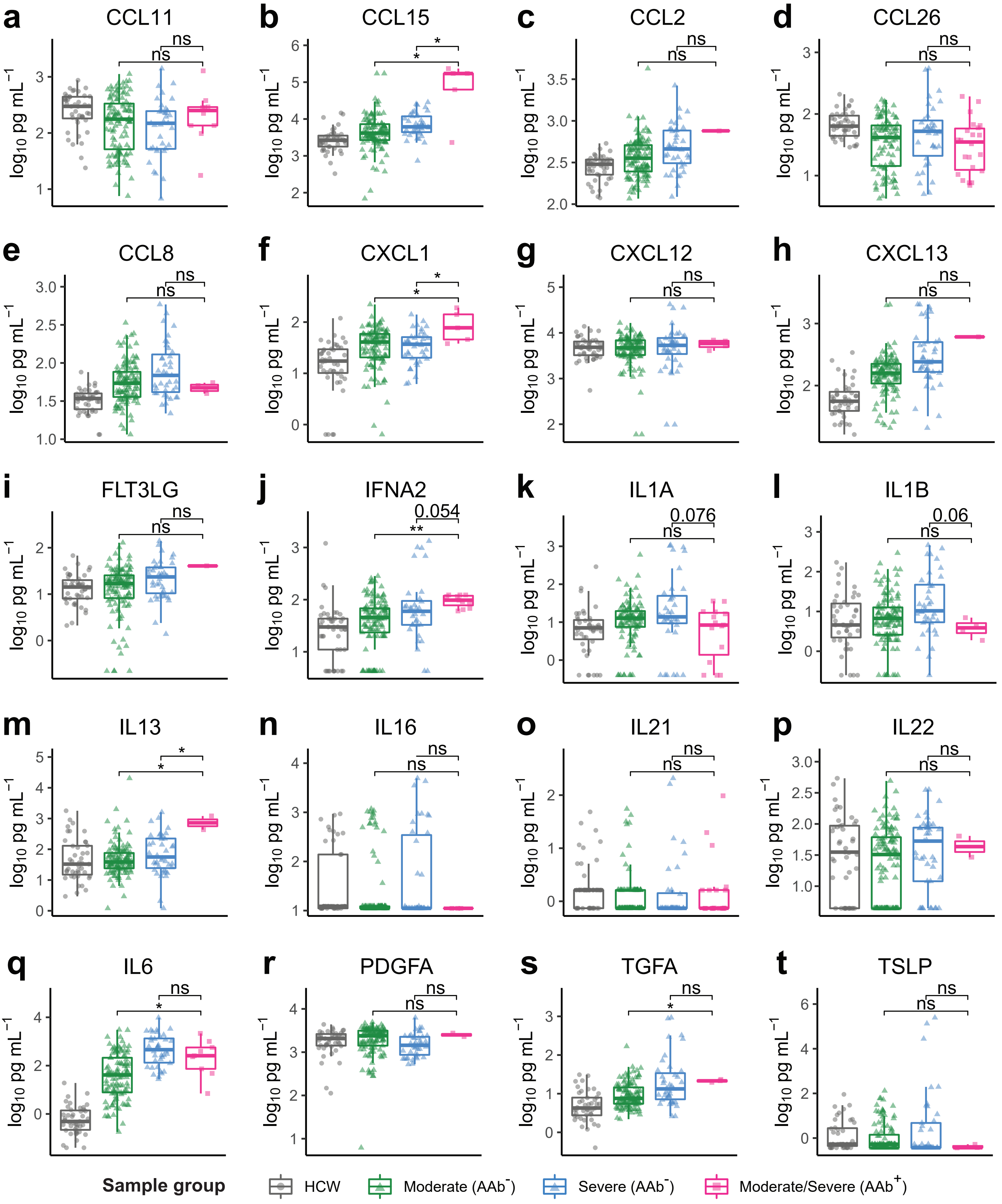


**Supplementary Figure 4. a-t,** Concentration of plasma CCL11 (**a**), CCL15 (**b**), CCL2 (**c**), CCL26 (**d**), CCL8 (**e**), CXCL1 (**f**), CXCL12 (**g**), CXCL13 (**h**), FLT3LG (**i**), IFNA2 (**j**), IL1A (**k**), IL1B (**l**), IL13 (**m**), IL16 (**n**) and IL21 (**o**), IL22 (**p**), IL6 (**q**), PDGFA (**r**), TGFA (**s**), and TSLP (**t**) measured by a Luminex assay in samples stratified by COVID-19 disease severity and REAP reactivity (AAb+; REAP score >= 2) against the corresponding cytokine. Significance was determined using two-sided, Wilcoxon rank-sum test; *P ≤ 0.05, **P ≤ 0.01, ***P ≤ 0.001, ****P ≤ 0.0001. All error bars in this figure represent standard deviation.

### Supplementary Figure 5

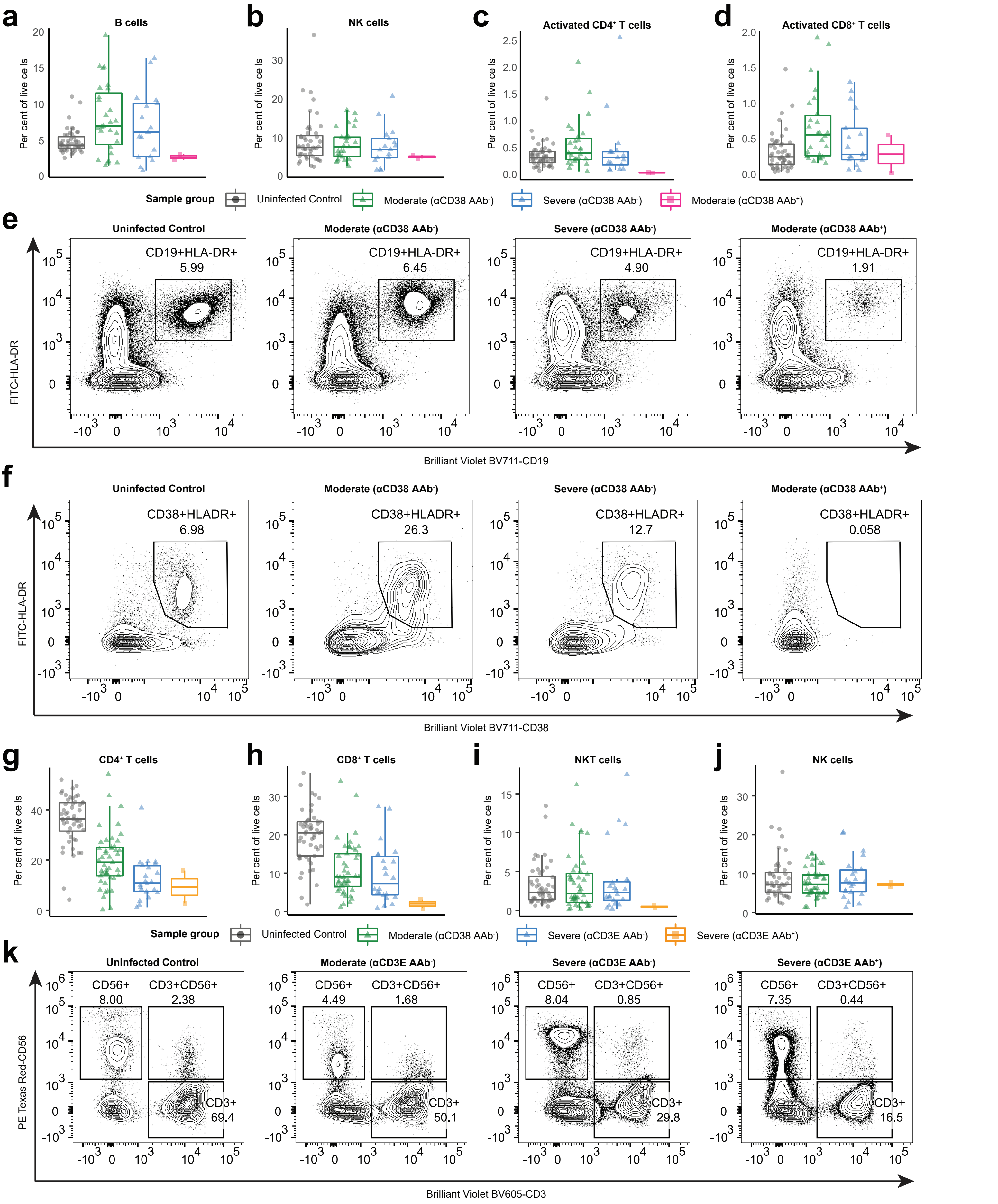


**Supplementary Figure 5. a-d,** Percent B cells (**a**), NK cells (**b**), Activated CD4+ T cells (**c**), activated CD8+ T cells (**d**) among peripheral leukocytes in samples stratified by COVID-19 disease severity and REAP reactivity (score >= 2) to CD38. **e,** Representative flow plot of B cells (CD19+HLA-DR+) for **a. f,** Representative flow plot of activated CD8+ T cells cells (CD19+HLA-DR+) for **d. g-j,** Percent CD4+ T cells (**g**), CD8+ T cells (**h**), NKT cells (**i**), and NK cells (**j**) among peripheral leukocytes in samples stratified by COVID-19 disease severity and REAP reactivity (score >= 2) to CD3E. **k,** Representative flow plot of T cells (CD3+), NK cells (CD56+), and NKT cells (CD3+CD56+) for **g-j**.

### Supplementary Figure 6

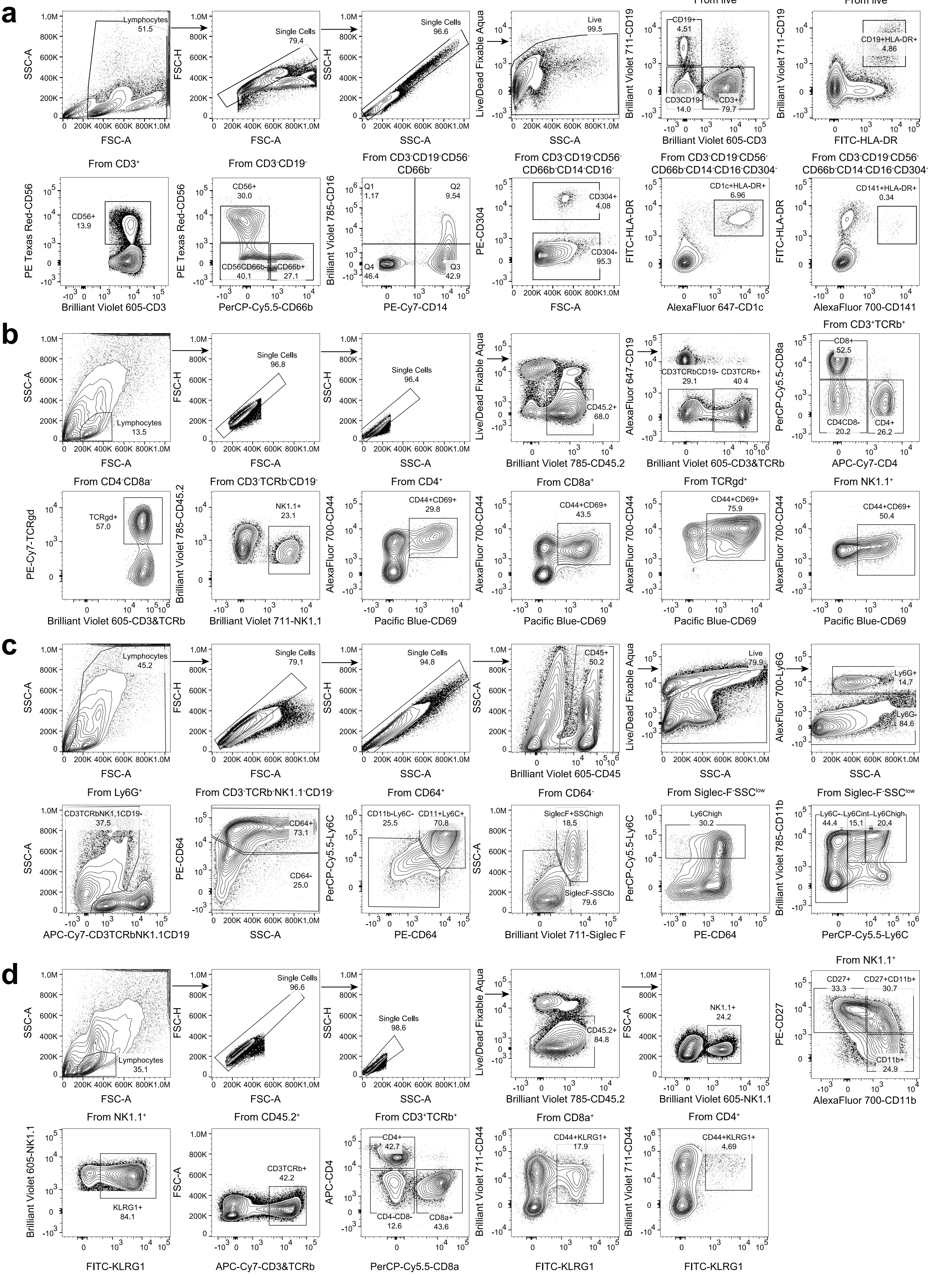

**Supplementary Figure 6. a,** Gating strategy to identify B cells in Fig. 2f, monocytes in **Fig. 2h-k**, T cells, NKT cells, and NK cells in **Supplementary Fig. 5.** in human PBMCs. **b,** Gating strategy to identify CD11b+Ly6Chigh monocytes and Ly6C+CD11b+CD64+ macrophages in mouse lung tissues described in **Fig. 3c-e. c,** Gating strategy to identify CD44+CD69+ lymphocytes in mouse lung tissues described in **Fig. 3f-g. d,** Gating strategy to identify KLRG1+ and CD11b+ NK cells described in **Fig. 3k-l.**

### Supplementary Figure 7

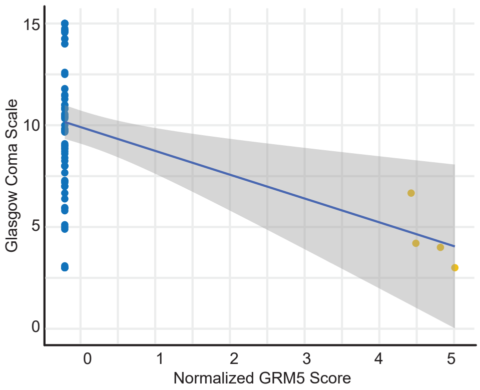


**Supplementary Figure 7.** Correlation of normalized GRM5 REAP scores with Glasgow Coma Scale scores in severe COVID-19 samples. Blue line shows a linear regression fit. Samples from the same patient were indicated with the same color points.
